# Topological data analysis communities reveal gene-environment-brain subtypes of major depression in UK Biobank and multi-site cohorts

**DOI:** 10.1101/2025.10.29.25339044

**Authors:** Emma Tassi, Alessandro Pigoni, Federica Colombo, Lidia Fortaner-Uyà, Cristina Colombo, Anna Maria Bianchi, Francesco Benedetti, Chiara Fabbri, Alessandro Serretti, GSRD network, Benedetta Vai, Paolo Brambilla, Eleonora Maggioni

## Abstract

Major depressive disorder (MDD) exhibits substantial clinical heterogeneity complicating prognosis definition and treatment selection. Characterizing MDD subtypes through distinct clinical manifestations could enhance personalized therapeutic approaches. We developed a topological data analysis (TDA) framework with graph-based community detection to identify homogeneous patient subgroups using multimodal data integration. We implemented a TDA pipeline in UK Biobank MDD participants with gene-environment (G-E, N=20,715) and gene-environment-neuroimaging (G-E-I, N=3,044) data. We systematically compared predictive capabilities across genetic, environmental, and neuroimaging features, alone and combined, for 18 health-related outcomes. For the best-predictive set of features identified for each outcome, a novel two-stage feature ranking approach identified features relevant for graph construction and community-based outcome differentiation. Cross-cohort validation utilized two independent datasets.

G-E interactions demonstrated superior predictive performance for 13 clinical outcomes, including treatment-resistant depression (TRD), symptom subtypes, and suicidal phenotypes. Community profiling revealed distinct vulnerability pathways: trauma-stress exposures linked to TRD and episode severity, while substance-behavioral profiles associated with anxious symptoms. Environmental factors emerged as primary determinants of most health outcomes, whereas neuroimaging features optimally predict medical comorbidities. Cross-cohort validation confirmed replication for multiple outcomes: self-harm behavior and anxious features (GSRD), TRD and vascular diseases (HSR), with consistent environmental stress-related predictive features across cohorts. TDA successfully identified clinically relevant MDD subgroups with unique multimodal signatures. These findings underscore the essential role of integrating genetic, environmental, and neuroimaging characteristics for robust health outcome prediction, establishing TDA-based community detection as an effective framework for MDD patient stratification and advancing precision medicine approaches in depression management.

**Significance Statement:** Topological Data Analysis (TDA) combined with community detection was used to identify clinically meaningful subgroups within Major Depressive Disorder (MDD) from multimodal UK Biobank data integrating genetic, environmental, and neuroimaging features. We systematically compared unimodal and multimodal feature sets to stratify patients across 18 health-related outcomes, with cross-cohort validation in independent datasets. Gene-by-environment interactions emerged as optimal predictors for mental health outcomes, revealing distinct vulnerability pathways: trauma-stress profiles predicted treatment resistance and episode severity, while substance-behavioral patterns were linked to anxious and neurovegetative symptoms. Brain imaging features best predicted medical comorbidities, particularly vascular diseases. Cross-cohort validation confirmed replication across populations. These findings establish TDA-based community detection as a powerful framework for MDD stratification, advancing precision psychiatry and personalized intervention strategies.

## Introduction

With over 300 million affected, major depressive disorder (MDD) is one of the world’s foremost causes of disability, placing a heavy burden on health systems (1–3). MDD is a complex disorder characterized by a high clinical heterogeneity, including a range of symptoms, illness courses, and response to treatment. This heterogeneity extends also to underlying etiologic mechanisms, such as genetics, neural substrates and environmental factors, and to comorbidities (4, 5). This broad variability complicates efforts to deliver optimal treatments, significantly hindering effective clinical management and patient-centered recovery paths (6).

When it comes to treatment, no antidepressant works for everyone, often requiring several trials: around one-third of individuals do not experience enough symptom relief after their initial antidepressant, and about 15% remain symptomatic even after multiple attempts (7, 8). Lack of response to at least two different antidepressant medications is usually defined as treatment-resistant depression (TRD), a condition with considerable personal and socioeconomic implications (9, 10).

This trial-and-error approach delays remission and considerably increases the overall burden of the disorder (11). Besides, life expectancy in patients with depressive disorders is usually shorter than the general population due to both suicide and natural causes of mortality (12). MDD is associated with vascular and cardiac problems (13), diabetes (14), autoimmune (15) and inflammatory disorders (16), and cancer (17). Finally, MDD increases the risk for suicidal behaviors (18), as mood disorders are the leading diagnoses associated with suicide attempts (19).

Therefore, one of the unmet needs in MDD management is the possibility to stratify patients, dissecting their heterogeneity in terms of clinical features, illness trajectories, and risk of developing negative outcomes.

Clinical studies that used depressive symptoms to characterize MDD subtypes have generally identified up to five different subtypes, including atypical, melancholic/typical, seasonal, psychotic, and mixed anxiety MDD (20, 21). Even with encouraging recent efforts to leverage multiple data sources for identifying MDD subtypes (e.g., symptomatology, genetic, biochemical, and neuroimaging data), these findings have shown substantial inconsistency (21, 22). Frequent limitations, such as insufficient validation and lack of replication, limiting their clinical relevance and applicability, generalizability as well as the mechanistic insights they yield (21, 23). The inability to adequately address this heterogeneity, particularly from a biological standpoint, hinders both drug discovery and the development of personalized treatments (8, 24). The classification of patients into homogeneous subgroups could facilitate our understanding of the underlying biological mechanisms (25) and the development of subtype-specific healthcare interventions, including individualized preventive and treatment strategies (8, 24).

Numerous data-driven methods for clinical stratification have been used to define MDD subgroups, such as k-means, tree-based clustering, and latent class analysis (26–29) (. Among the emerging data-driven methodologies, topological data analysis (TDA) has proven to excel at handling high-dimensional, nonlinear datasets, identifying patterns across different scales, and remaining stable despite data variations (30, 31). This allows to overcome the limitations faced by traditional clustering approaches when data shapes become overly complex, as is often the case with neuropsychiatric disorders (32, 33). TDA employs methods based on algebraic topology, defined as the area of mathematics that focuses on the notion of shape and connectivity, and attempts to study patterns (i.e., features) within the data. Specifically, TDA investigates the “shape” (i.e., underlying topology) of the data, making it ideal for uncovering latent topological structures (34, 35). TDA can effectively manage higher-order interactions, which are common in neuropsychological and brain imaging data, and provides intuitive outputs for the exploration of the dataset, allowing researchers to see how features converge or diverge within defined clusters (36).

TDA has been successfully employed in different medicine fields (30, 37), including neuropsychiatric disorders such as autism-spectrum disorders (33) and schizophrenia (38). One of the available approaches, namely TDA Mapper, produces a graph structure where nodes represent overlapping subsets of patients and edges encode similarity relations, enabling the integration with graph-based community detection methods. This approach leverages community detection algorithms, which aim to maximize modularity and identify patient "communities" or subgroups that are densely interconnected but sparsely connected to the rest of the graph (39, 40). Recent studies have increasingly applied graph-based community detection to various types of biological data to uncover subgroups characterized by distinct phenotypes or biological pathways. These approaches were successfully implemented across neuroimaging datasets (41–45), clinical measurements, and molecular networks (46) to identify meaningful communities. However, systematic comparisons of different feature sets in terms of community-based outcome profiling and comprehensive replication across independent cohorts are lacking, limiting our understanding of the generalizability and clinical utility of TDA-derived communities for managing complex psychiatric disorders like MDD.

The present study addresses these gaps by developing and applying a novel TDA-based framework for graph-based community detection and prediction of clinical outcomes in MDD. Training was performed using multimodal genetic, environmental, and neuroimaging data from the UK Biobank (UKB) subsample affected with MDD, whereas testing was performed using data from two independent MDD cohorts. This framework, for the first time, systematically compares community-based outcome prediction across multimodal feature sets, applies multiple community detection algorithms, and characterizes clinically meaningful subgroups through comprehensive testing across multiple health-related outcomes. Our approach introduces a novel feature ranking pipeline and tests the translation potential of findings across two independent MDD cohorts, providing the first systematic replication of TDA-derived communities in psychiatric research.

This study builds upon the multimodal TDA research framework established by Tassi et al. (2025) (47), which explored gene-environment-brain topology for clinical outcome prediction in MDD using UKB data. Leveraging the same large-scale multimodal dataset from the UKB, our study aimed to extend previous SAFE score-based outcome prediction findings by shifting from a prediction-focused approach to a patient stratification perspective using a graph-based community detection algorithm.

The previous TDA application provided valuable insights into the differential predictive capabilities of genetic, environmental, and neuroimaging predictive feature sets for various clinical outcomes, establishing which multimodal combinations optimally predicted different aspects of MDD presentation. While this outcome-focused methodology successfully identified predictive relationships within TDA networks, it did not address patient stratification into homogeneous clinical subgroups. Building on the previous methodology, the present study takes a complementary step forward by implementing community detection algorithms applied on TDA graph to cluster a large MDD sample. Specifically, our approach aims to identify homogeneous MDD patient communities with distinct multimodal risk profiles and evaluate the replicability of these findings across independent cohorts.

Based on the established effectiveness of multimodal TDA frameworks and the clinical need for patient stratification, we hypothesized that: (1) TDA-based community detection will reveal distinct MDD subgroups with specific multimodal signatures linked with health outcomes; (2) gene-imaging-environment interactions will play a key role in predicting mental and general health outcomes; and (3) identified communities will demonstrate clinically meaningful risk stratification patterns with good replicability across independent cohorts, providing a robust framework for personalized MDD management.

## Results

### 2.1 Participants characterization

We analyzed three MDD cohorts: UK Biobank (UKB), the European Group for the Study of treatment Resistant Depression (GSRD, N=1,017), and a cohort from Hospital San Raffaele (HSR, N=87). The characteristics of UKB and replication samples are detailed in **Table 1**.

**Table 1.**
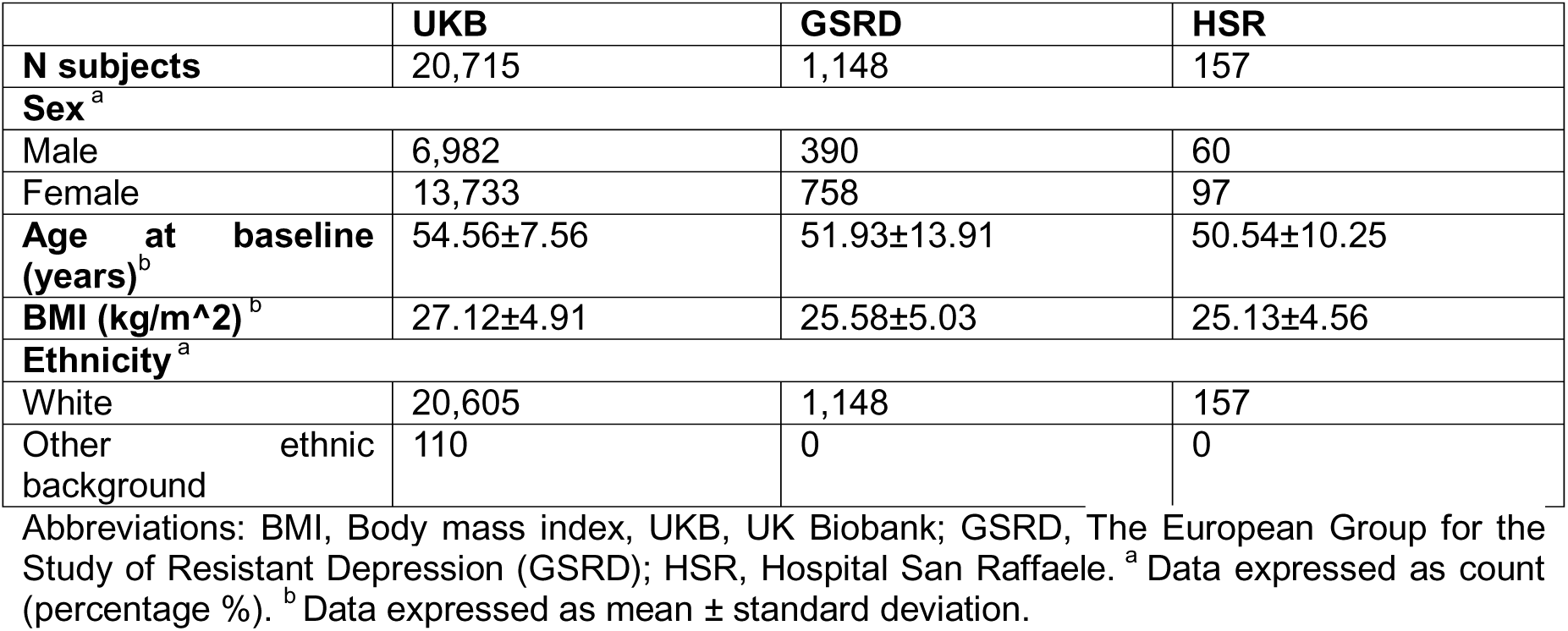
Main clinical and socio-demographic characteristics of the UKB sample and independent sets of GSRD and HSR.

|  | **UKB** | **GSRD** | **HSR** |
| --- | --- | --- | --- |
| **N subjects** | 20,715 | 1,148 | 157 |
| **Sex<sup>a</sup>** |  |  |  |
| Male | 6,982 | 390 | 60 |
| Female | 13,733 | 758 | 97 |
| **Age at baseline (years)<sup>b</sup>** | 54.56±7.56 | 51.93±13.91 | 50.54±10.25 |
| **BMI (kg/m<sup>2</sup>)<sup>b</sup>** | 27.12±4.91 | 25.58±5.03 | 25.13±4.56 |
| **Ethnicity<sup>a</sup>** |  |  |  |
| White | 20,605 | 1,148 | 157 |
| Other ethnic background | 110 | 0 | 0 |

Abbreviations: BMI, Body mass index, UKB, UK Biobank; GSRD, The European Group for the Study of Resistant Depression (GSRD); HSR, Hospital San Raffaele. <sup>a</sup> Data expressed as count (percentage %). <sup>b</sup> Data expressed as mean ± standard deviation.

In UKB, MDD participants with available gene-environment (G-E) data (N=20,715) and a subsample with gene-environment-imaging (G-E-I) data (N=3,044) were considered. The GSRD sample (N=1,017) was taken entirely with available G-E features, whereas for the HSR sample N=87 participants had G-E features, and N=71 had G-E-I features. Different subsets were extracted from each cohort based on the availability of information on the 18 health-related outcomes of interest, spanning depression severity measures, TRD, psychosocial functioning, and medical comorbidities (see **Table 2**).

**Table 2.** Description of clinically relevant outcomes considered in UKB sample and independent sets.

| UKB, O=18 | GSRD, O=6 | HSR, O=11 |
| --- | --- | --- |
| • TRD<br>• Depression with atypical neurovegetative symptoms<br>• Depression with anxious features<br>• Number of depression episodes<br>• Health satisfaction<br>• Family relationship satisfaction<br>• Friendship relationship satisfaction<br>• Work job satisfaction<br>• Duration of worst depression<br>• Frequency of depressed days during worst episode of depression<br>• Impact on normal roles during worst period of depression<br>• Thoughts of death during worst depression<br>• Ever self-harmed<br>• Belief that owns life is meaningful<br>• Depression possibly related to stressful or traumatic event<br>• Cancer diagnosed by doctor<br>• Diabetes diagnosed by doctor<br>• Vascular heart problems | • TRD<br>• Depression with anxious features<br>• Duration of current depressive episodes<br>• Thoughts of death during current depressive episodes<br>• Ever self-harmed<br>• Somatic disorders (i.e., include diabetes, cancer and vascular diseases) | • TRD<br>• Depression with atypical neurovegetative symptoms<br>• Depression with anxious features<br>• Number of depression episodes<br>• Duration of current depressive episodes<br>• Thoughts of death during current depressive episodes<br>• Ever self-harmed<br>• Belief that owns life is meaningful<br>• Cancer diagnosed by doctor<br>• Diabetes diagnosed by doctor<br>• Vascular heart problems |

Abbreviations: O, outcome; TRD, treatment-resistant depression.

### 2.2 Pipeline Overview

The TDA and community detection analysis workflow, schematized in **Figure 1**, had two main steps: (1) identifying the most predictive feature sets -among gene, environment, and brain imaging ones, alone and combined-for stratification of health outcomes in the UKB subsample with MDD, and (2) testing their cross-cohort generalizability in two independent MDD datasets. We employed multimodal genetic, environmental, and neuroimaging feature sets as input for the TDA Mapper to assess their differential capability to cluster MDD individuals into communities with distinct health outcome profiles. Multimodal neuroimaging predictors included: T1-weighted structural MRI (sMRI), diffusion weighted MRI (dMRI), resting-state functional MRI (rs-fMRI), and task-based functional MRI (t-fMRI) data from emotional processing paradigms. Genetic predictors consisted of polygenic risk scores (PRSs) for psychiatric disorders, behavioral traits, and immune-cardiometabolic conditions. Environmental predictors encompassed lifetime experiences including childhood trauma, adult stress, substance use patterns, and social support factors.

**Figure 1.**
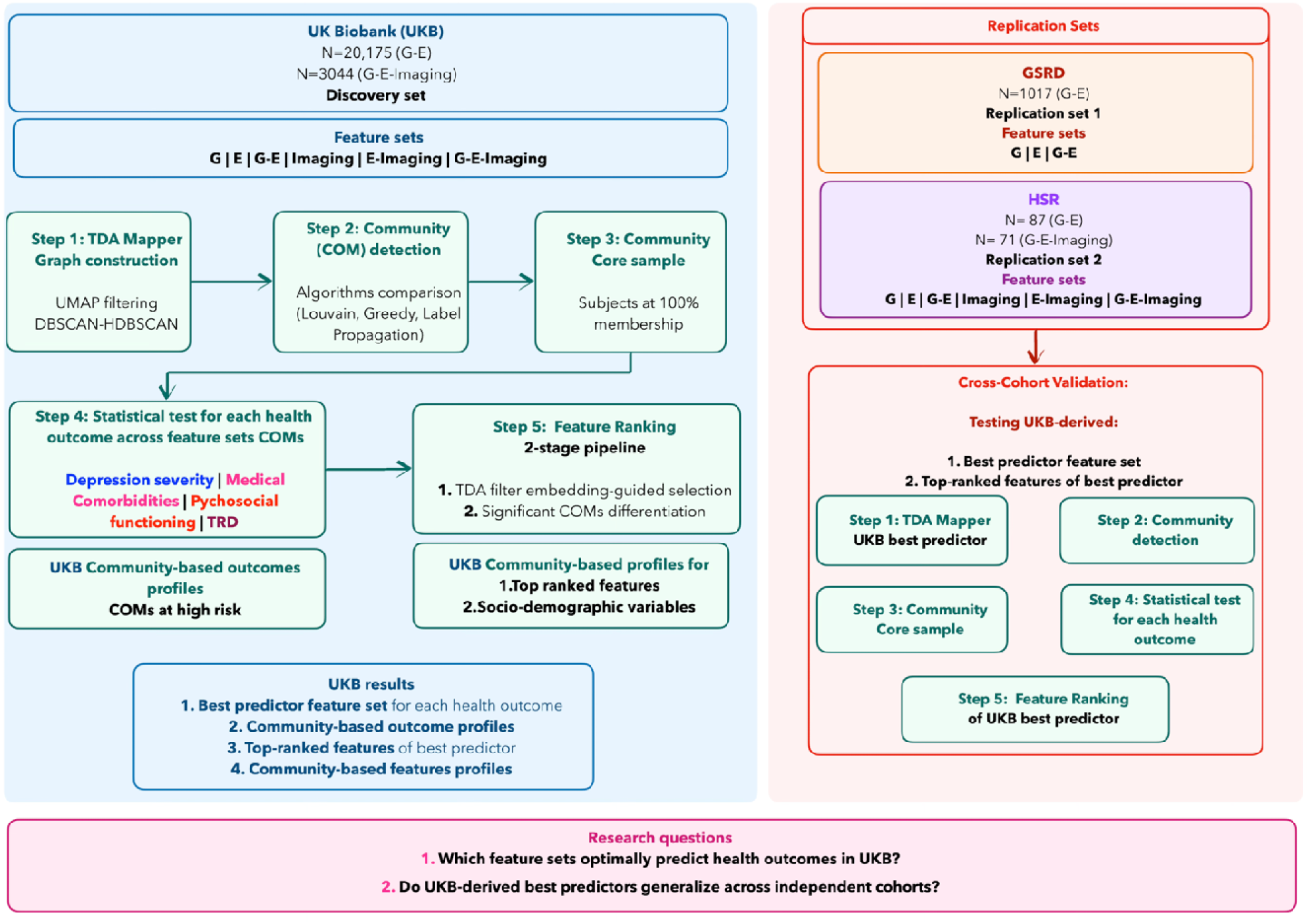
TDA-based data analysis workflow for MDD patient stratification. Five-step TDA framework applied to UK Biobank (UKB) discovery dataset (left panel) and validated across two independent replication cohorts (right panel, GSRD and HSR), addressing: (1) identifying optimal predictive feature sets for health outcomes stratification, and (2) testing cross-cohort generalizability of UKB-derived best predictors. (Left panel) **Step 1: TDA Mapper** constructs graphs using UMAP dimensionality reduction and density-based clustering across six multimodal feature sets (G, genetic; E, environment; G-E, genetic-environment; E-Imaging, environment-imaging; G-E-Imaging, genetic-environmental-imaging). **Step 2: Community Detection** applies algorithms selected based on modularity and performance metrics. **Step 3: Core Sample Extraction** retains subjects with 100% membership confidence. **Step 4: Community-based Outcome Stratification** performs statistical testing across 18 health outcomes, identifying best predictors based on statistical significance and effect size. **Step 5: Feature Ranking** implements a two-stage pipeline combining UMAP-guided selection with community-based validation. (Right panel) For cross-cohort validation, the complete pipeline is applied to replication datasets using UKB-derived best predictors to assess generalizability across independent populations.

The analysis was implemented through a systematic five-step pipeline (Methods) in the UKB discovery dataset (48): (1) TDA Mapper application to construct graph structures where nodes represent patient clusters and edges encode similarity relationships; (2) Community detection using algorithms selected based on modularity and network performance metrics to partition graphs into meaningful patient subgroups; (3) Core sample extraction retaining subjects with 100% membership confidence to a single community to ensure reliable community assignments; (4) Community-based outcome stratification performing statistical testing across the 18 health outcomes to identify best predictive feature sets; (5) Feature ranking implementing a novel two-stage pipeline combining UMAP-guided dimensionality reduction with community-based validation to identify features most influential for graph construction and outcome differentiation.

For the UKB discovery dataset, all six feature sets (G, E, G-E, Imaging, E-Imaging, G-E-Imaging) were systematically compared. For replication cohorts (GSRD and HSR), the complete pipeline was applied using UKB-derived best predictors to assess cross-cohort generalizability and feature consistency across independent MDD populations. Specifically, we applied steps 1-4 of the pipeline using only the UKB-identified best predictor feature set for each outcome, followed by feature ranking analysis (step 5) to assess cross-cohort feature consistency and replicability. Details on data preprocessing, TDA parameters, and statistical methods are provided in Methods sections.

### 2.3 TDA Graph structural characteristics

For UKB, TDA graphs were obtained on G-E datasets (G, E, G-E; 20,715 subjects) and imaging-based datasets (G-E-Imaging, E-Imaging, Imaging; 3,044 subjects). The TDA graphs’ structures obtained on UKB feature sets can be visualized in **Figure 2**. Among the community detection algorithms tested, Louvain modularity showed the highest effectiveness in detecting communities across all UKB feature sets, evaluated in terms of coverage, performance and modularity. The G-E-Imaging, E-Imaging, Imaging datasets generated 4 robust communities each with core sample sizes ranging from 130 to 468 subjects and modularity in the [0.43-0.46] range, while larger G, E, G-E datasets showed 4 robust communities with more variable core sample distributions (from 51 to 8,287 subjects) and modularity reaching 0.60 for the E feature set. Detailed TDA graph characteristics, community detection algorithm performance, and core sample sizes for each community obtained on UKB feature sets are provided in **Table S10.**

**Figure 2.**
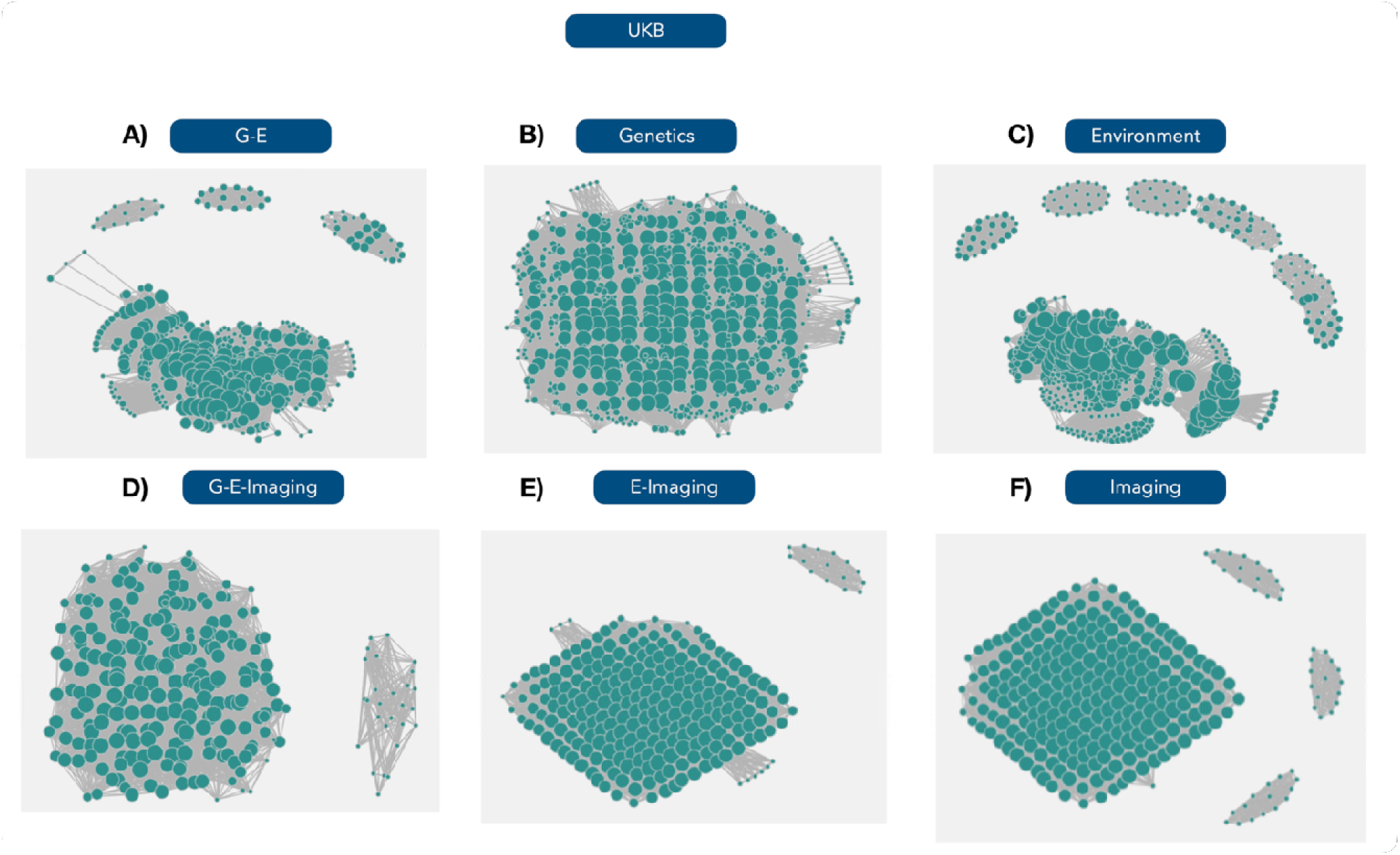
TDA graph structures for all predictors tested in the UKB dataset. including: G-E, Genetics, Environment, G-E-Imaging, E-Imaging, Imaging feature sets. Each visualization displays the network topology obtained where nodes represent clustered subjects with similar feature profiles and edges connect overlapping patient neighbourhoods.

For the replication sets, Louvain algorithm showed the highest effectiveness in detecting communities in terms of modularity, coverage, and performance on all feature sets of GSRD. In HSR, Louvain algorithm was selected for community-detection of G, E, G-E, and G-E-Imaging, while the Greedy algorithm led to optimal performances for E-Imaging and Imaging sets. GSRD achieved exceptionally high modularity (0.97) for E features and generated from 3 to 13 robust communities across feature sets with core sample sizes ranging from 29 to 255 subjects. HSR reached the highest modularity for the E-Imaging (0.65) datasets and across feature sets produced 2-4 robust communities with smaller core sample sizes (from 6 to 28 subjects) due to the limited sample size. Comprehensive details on TDA graph characteristics, community detection algorithm performance, and core sample sizes for each community obtained from replication sets can be found in **Tables S11** and **S12**.

### 2.4 Community-based outcome prediction

#### UKB dataset

For the UKB dataset, statistics and effect sizes for all outcomes and feature sets’ communities are reported in Table 4. G-E communities emerged as best predictors for the majority of outcomes, showing the strongest prediction for 13 out of 18 health-related outcomes, including TRD, depression subtypes (atypical neurovegetative and anxious features), functional outcomes (number of depression episodes, health, family relationship, and friendship satisfaction), and suicidal ideation or behaviors (thoughts of death and self-harm). Domain-specific best predictors emerged for medical comorbidities: G-E-Imaging for depression related to stressful events, E-Imaging for cancer diagnosis, Imaging for vascular problems, and G for diabetes.

**Table 3.** Genetic, environmental and imaging characteristics included in UKB sample and independent sets.

| **Environment** |  |  |
| --- | --- | --- |
| **UKB (P=33)** | **GSRD (P=5)** | **HSR (P=12)** |
| • Felt love as a child<br>• Physically abused by family as a child<br>• Felt hated by family member as a child<br>• Sexually molested as a child<br>• Someone to take to doctor when needed as a child<br>• Been in a confiding relationship as an adult<br>• Physical violence by partner or ex-partner as an adult<br>• Belittlement by partner or ex-partner as an adult<br>• Sexual interference by partner or ex-partner without consent as an adult<br>• Able to pay rent/mortgage as an adult<br>• Victim of sexual assault<br>• Victim of physically violent crime<br>• Been in serious accident believed to be life-threatening<br>• Witnessed sudden violent death<br>• Diagnosed with life-threatening illness<br>• Been involved in combat or exposed to war-zone<br>• Alcohol intake frequency<br>• Ever Taken Cannabis<br>• Childhood stressful events<br>• Adulthood stress<br>• Serious illness, injury, assault to yourself or assault of a close relative in the last 2 years<br>• Death of a close relative or death of a spouse or partner in the last 2 years<br>• Stress related to marital separation/divorce or to financial difficulties in the last 2 years<br>• Smoking status<br>• Major dietary changes because of illness in the last 5 years<br>• Major dietary changes because other reason in the last 5 year | • Social activities (item 2 of the Sheenan Disability Scale)<br>• Marital status<br>• Working status<br>• Alcohol intake frequency<br>• Smoking status | • Felt love as a child<br>• Physically abused by family as a child<br>• Felt hated by family member as a child<br>• Sexually molested as a child<br>• Someone to take to doctor when needed as a child<br>• Alcohol intake frequency<br>• Serious illness, injury, assault to yourself or assault of a close relative in the last 3 years<br>• Death of a close relative or death of a spouse or partner in the last 3 years<br>• Stress related to marital separation/divorce or to financial difficulties in the last 3 years<br>• Smoking status<br>• Major dietary changes because other reason in the last 3 years |
| • Frequency of friend / family visits<br>• Leisure / social activities<br>• Able to confide<br>• Morning/evening person (chronotype)<br>• Summed MET minutes per week for all activity<br>• Z adjusted T/S log (i.e., Telomeres length)<br>• Neuroticism-EPQ RS |  |  |
| **Genetics (PRS)** |  |  |
| **UKB (P=20)** | **GSRD (P=18)** | **HSR (P=19)** |
| • BD<br>• Schizophrenia<br>• MDD<br>• Anorexia nervosa<br>• Autism<br>• ADHD<br>• Alcohol dependence<br>• Neuroticism<br>• Smoking<br>• Alcohol consumption<br>• Years of education<br>• College<br>• C-reactive protein<br>• Glycated hemoglobin<br>• Triglycerides<br>• LDL cholesterol<br>• HDL cholesterol<br>• Coronary artery disease<br>• BMI<br>• Type 2 diabetes mellitus | • BD<br>• Schizophrenia<br>• MDD<br>• Anorexia Nervosa<br>• Autism<br>• ADHD<br>• Alcohol dependence<br>• Smoking (N sigarette / day)<br>• Alcohol consumption (daily)<br>• Years of education<br>• C-reactive protein<br>• Glycated hemoglobin<br>• Triglycerides<br>• LDL cholesterol<br>• HDL cholesterol<br>• Coronary artery disease<br>• BMI<br>• Type 2 diabetes mellitus | • BD<br>• Schizophrenia<br>• MDD<br>• Anorexia Nervosa<br>• Autism<br>• ADHD<br>• Alcohol dependence<br>• Neuroticism<br>• Smoking (N sigarette / day)<br>• Alcohol consumption (daily)<br>• Years of education<br>• C-reactive protein<br>• Glycated hemoglobin<br>• Triglycerides<br>• LDL cholesterol<br>• HDL cholesterol<br>• Coronary artery disease<br>• BMI<br>• Type 2 diabetes mellitus |
| **Imaging** |  |  |
| **UKB (P=606)** | **GSRD (/)** | **HSR (P=344)** |
| • sMRI, P=200<br>14 Subcortical Volumes, 62 GMV (Desikan-Killiany atlas), 62 CT (Desikan-Killiany atlas), 62 SA (Desikan-Killiany atlas)<br>• dMRI, P=288<br>48 FA ROIs, 48 ODI, 48 ICVF, 48 ISOVF, 48 RD, 48 L1 directions, all from JHU atlas<br>• rs-fMRI, P=110<br>55 strengths of positive weights, 55 strengths of negative weights<br>• t-fMRI, P=8<br>4 measures of 90th percentile of BOLD effect, 4 measures z-statistics for faces-shapes contrast, faces activation, and shapes activation in group-defined mask, and for faces-shapes contrast in group-defined amygdala activation mask. | / | • sMRI, P=200<br>14 Subcortical Volumes, 62 GMV (Desikan-Killiany atlas), 62 CT (Desikan-Killiany atlas), 62 SA (Desikan-Killiany atlas)<br>• dMRI, P=144<br>48 FA ROIs, 48 RD, 48 L1 directions, all from JHU atlas |

Abbreviations: MET, metabolic equivalent task; EPQ- RS, Eysenck Personality Questionnaire – Revised; PRS, Polygenetic risk scores; sMRI, structural MRI; rs-fMRI, resting-state functional MRI; t-fMRI, task-based functional MRI; dMRI, diffusion weighted MRI; JHU atlas, Johns Hopkins University atlas; GMV, gray matter volume; CT, cortical thickness; SA, surface area; FA, Fractional Anisotropy; RD, radial diffusivity; ODI, orientation dispersion index; ICVF, Intracellular Volume Fraction; ISOVF, Isotropic Compartment Volume Fraction; L1, L1 directions; PRS, Polygenetic risk scores; BD, Bipolar disorder; MDD, Major depressive disorder; ADHD, attention deficit hyperactivity disorder; LDL, Low-Density Lipoprotein cholesterol; BMI, body mass index; HDL, High-Density Lipoprotein cholesterol.

**Table 4.** Statistics and effect sizes extracted for each outcome across communities obtained on each feature set. The best predictor was identified as the feature set showing the greatest statistical significance across communities (p<0.05, corrected for each outcome across the number of feature sets tested, N=6). Effect sizes calculated using Eta² for variables tested with KW and Cramer’s V for variables tested with Chi2 tests are reported. P-values reported in the table are not corrected for Bonferroni. P-values and statistics reported in bold are associated with the best predictors.

|  | **G-E-**<br>**Imaging** | **E-Imaging** | **Imaging** | **Environment** | **G-E** | **Genetics** |
| --- | --- | --- | --- | --- | --- | --- |
| TRD | NS | NS | NS | NS | **p=3.85e-04**<br>**(Chi2: 18,**<br>**Cramer V:**<br>**0.12)** | NS |
| Depression with atypical neurovegetative symptoms | NS | NS | NS | p=0.00130<br>(Chi2: 15,<br>Cramer V:<br>0.04) | **p=6.11e-08**<br>**(Chi2: 36.4,**<br>**Cramer V:**<br>**0.1)** | NS |
| Depression anxious features | NS | NS | NS | NS | **p=6.83e-07**<br>**(Chi2:**<br>**31.45, Cramer**<br>**V: 0.1)** | NS |
| N depression episodes | NS | NS | NS | p=1.45e-10<br>(Hstat: 48, $\eta^2$:<br>0.01) | **p=1.97e-16**<br>**(Hstat: 76.23,**<br>**$\eta^2$: 0.03)** | p<0.001,<br>p=2.22e-04<br>(Hstat: 5.70,<br>$\eta^2$: 0.0019) |
| Health Satisfaction | NS | NS | NS | p=0.003<br>(Hstat: 13, $\eta^2$:<br>0.0025) | **p=1.94e-07**<br>**(Hstat: 34, $\eta^2$:**<br>**0.014 )** | NS |
| Family relationship satisfaction | p=0.006<br>(Hstat: 12,<br>$\eta^2$: 0.03) | NS | NS | p=2.1e-07<br>(Hstat: 34, $\eta^2$:<br>0.007) | **p=4.62e-19**<br>**(Hstat: 88, $\eta^2$:**<br>**0.041)** | NS |
| Friendship relationship satisfaction | p=0.002<br>(Hstat: 15,<br>$\eta^2$: 0.04) | NS | p=0.04<br>(Hstat: 11,<br>$\eta^2$: 0.025) | p=0.04<br>(Hstat: 8, $\eta^2$:<br>0.0013) | **p=1.40e-06**<br>**(Hstat: 30, $\eta^2$:**<br>**0.012)** | NS |
| Work Job Satisfaction | NS | NS | NS | NS | NS | NS |
| Duration Worst Depression | p=0.00034<br>(Hstat: 18.5,<br>$\eta^2$: 0.017) | p=0.0003<br>(Hstat: 19,<br>$\eta^2$: 0.017) | p=0.00032<br>(Hstat: 18.6,<br>$\eta^2$: 0.016) | p=0.02<br>(Hstat: 9, $\eta^2$:<br>0.0006) | **p=1.45e-26**<br>**(Hstat: 123,**<br>**$\eta^2$: 0.02)** | NS |
| Frequency of depressed days | NS | p=0.01<br>(Hstat: 10,<br>H<sup>2</sup>: 0.008) | p=0.02<br>(Hstat: 9,<br>$\eta^2$: 0.0062) | p=0.0075<br>(Hstat: 12, H<sup>2</sup>:<br>0.0008) | **p=8.02e-06**<br>**(Hstat: 26, $\eta^2$:**<br>**0.004)** | NS |
| Impact of depression on normal roles | NS | NS | NS | p=0.0061<br>(Hstat: 12, $\eta^2$:<br>0.0008) | **p=6.39e-11**<br>**(Hstat: 50, $\eta^2$:**<br>**0.008)** | NS |
| Thoughts of death | NS | p=0.03<br>(Chi2: 8,<br>CramerV:<br>0.09) | p=0.01<br>(Chi2: 16,<br>CramerV:<br>0.12) | p=8.80e-06<br>(Chi2: 26,<br>Cramer V:<br>0.04) | **p=4.35e-15**<br>**(Chi2:**<br>**70, Cramer V:**<br>**0.11)** | NS |
| Ever self harmed | p=0.03<br>(Chi2:17,<br>Cramer V:<br>0.009) | NS | NS | p=8.27e-34<br>(Chi2: 156,<br>Cramer V:<br>0.12) | **p=1.51e-37**<br>**(Chi2:**<br>**174, Cramer**<br>**V: 0.17)** | p=2.56e-04<br>(Chi2: 19,<br>Cramer V:<br>0.07) |
| Belief own life is meaningful | NS | NS | NS | p=1.46e-13<br>(Hstat: 63, $\eta^2$:<br>0.005) | **p=4.16e-48**<br>**(Hstat: 223,**<br>**$\eta^2$: 0.04)** | NS |
| Depression related to stressful or traumatic events | **p= 5.90e-04**<br>**(Chi2: 17,**<br>**Cramer V:**<br>**0.14)** | p=0.01<br>(Chi2: 11,<br>Cramer V:<br>0.11) | p=0.0039<br>(Chi2:13,<br>Cramer V:<br>0.11) | NS | NS | NS |
| Cancer | NS | **p=9.51e-06**<br>**(Chi2:**<br>**13, Cramer**<br>**V: 0.11)** | NS | NS | NS | NS |
| Diabetes | NS | p=0.008<br>(Chi2: 12,<br>Cramer:<br>0.10) | NS | p=0.006,<br>(Chi2: 12,<br>Cramer V:<br>0.03) | p=0.003<br>(Chi2:<br>13, Cramer V:<br>0.04) | **p=2.12e-04**<br>**(Chi2:**<br>**19.5, Cramer**<br>**V: 0.07)** |
| Vascular Problems | p=0.03<br>(Hstat: 8.6,<br>$\eta^2$: 0.005) | **p=7.60e-10**<br>**(Hstat:**<br>**45, $\eta^2$:**<br>**0.041)** | **p=6.32e-10**<br>**(Hstat:**<br>**46, $\eta^2$:**<br>**0.044)** | NS | 3.185e-06<br>(Hstat: 28, $\eta^2$:<br>0.004) | p=2.31e-06<br>(Hstat: 28,<br>$\eta^2$: 0.006) |

Abbreviations: G-E-Imaging, genetic-environment; G-E, genetic-environment; TRD, treatment resistant depression; N, number of; Hstat, statistics from KW test; Chi2 statistics from Chi2 test; $\eta^2$ , effect size from Cohen's calculation; Cramer V, effect size from Cramèr calculation; NS, not significant.

Community-based outcome risk ranking analysis revealed distinct vulnerability patterns (**Table S13**). For G-E features, both community n.1 (COM1) and community n.4 (COM4) emerged as highest-risk groups for severe depression presentations, with partially different profiles. COM4 was characterized by high-risk group for depression with anxious and atypical neurovegetative features, daily depressive symptoms, suicidal ideation, and self-harm behaviours. COM1 showed the highest risk for TRD, depression episode severity, and stress-related depression vulnerability. Community n.2 (COM2) and community n.3 (COM3) showed elevated risk for outcomes related to wellbeing, with COM2 exhibiting the highest health and friendship relationship dissatisfaction, while COM3 displayed the greatest family relationship dissatisfaction and functional impairment. For other predictor sets, COM1 on G-E-Imaging showed highest stress-related depression risk, COM2 on E-Imaging demonstrated elevated cancer risk, COM2 on Imaging showed the highest vascular problems risk, and COM3 on G exhibited the highest diabetes risk.

#### Replication datasets

We tested whether the UKB-derived best predictor feature sets could successfully differentiate outcome subgroups in independent samples.

In GSRD, the best predictors identified on UKB were significantly replicated for two outcomes, which were significantly different across the relative communities (p<0.05): G-E predictor for self-harm behaviour (p=0.013, Cramer’s V: 0.12) and depression with anxious features (p=0.041, Cramer’s V: 0.10). Other G-E-predicted outcomes showed trend-level significance: TRD (p=0.057, Cramer’s V: 0.09) and medical comorbidities (p=0.075, Cramer’s V: 0.09), while thoughts of death showed no replication. In HSR, two feature set-outcome pairs demonstrated significant replication (p<0.05): G-E predictor for TRD (p=0.03, Cramer’s V: 0.44) and Imaging predictor for vascular problems (p=0.013, Cramer’s V: 0.39). The TDA graphs’ structures for these successfully replicated UKB-derived best predictors in independent cohorts are illustrated in **Figure S1**. In summary, G-E features demonstrated robust cross-cohort predictive capability for self-harm behaviour and anxious depression features in GSRD, and for TRD in HSR, while Imaging features replicated for vascular problems in HSR, supporting transferability of these optimal predictive relationships across independent cohorts.

### 2.5 Feature ranking and community characterization

#### UKB dataset

The key features for graph construction and community differentiation in UKB best predictive sets are shown in **Table 5**. The UMAP embedding correlations for features are presented in **Figure S2-S5**, showing the relationship between individual features and the low-dimensional topological space.

**Table 5.** Ranking of the most influential characteristics driving community structure from the best predictive feature set for all outcomes for UKB sample. The p-value significance threshold (Bonferroni corrected) associated with each features set is reported in the table.

| Predictor | Outcomes for which feature set is best predictor | Top-Ranked Features |
| --- | --- | --- |
| G-E-Imaging | O=1<br>Depression related to stressful or traumatic events | **461 features with significant corrected correlation with UMAP embedding; Top 20 features:**<br>Area of Superiorfrontal - Right<br>Area of Superiorfrontal - Left<br>Area of Superiortemporal - Right<br>Mean RD in SuperiorCoronaRadiata on FA skeleton - Left<br>Area of Superiortemporal - Left<br>Area of Lateralorbitofrontal - Left<br>Mean RD in SuperiorCoronaRadiata on FA skeleton - Right<br>Mean RD in PosteriorCoronaRadiata on FA skeleton - Left<br>Area of Lateralorbitofrontal - Right<br>Mean RD in PosteriorCoronaRadiata on FA skeleton - Right<br>Mean RD in SuperiorLongitudinalFasciculus on FA skeleton - Right<br>Area of Medialorbitofrontal - Left<br>Mean RD in GenuOfCorpusCallosum on FA skeleton_Instance2<br>Mean RD in SuperiorLongitudinalFasciculus on FA skeleton - Leftstan<br>VolumeOfLateralorbitofrontal - Right<br>Area of Middletemporal - Right<br>Mean RD in SagittalStratum on FA skeleton - Right<br>Mean RD in SuperiorFronto_occipitalFasciculus on FA skeleton - Left<br>VolumeOfMiddletemporal - Left<br>Mean RD in SuperiorFronto_occipitalFasciculus on FA skeleton - Right |
| **E-Imaging** | O =1<br>Cancer diagnosed | **441 features with significant corrected correlation with UMAP embedding; Top 20 features:**<br>Area of Superiorfrontal - Left<br>Area of Rostralanteriorcingulate - Left<br>Area of Middletemporal - Right<br>Area of Precentral - Left<br>Area of Precentral - Right<br>Area of Postcentral - Left<br>Area of Postcentral - Right<br>Area of Medialorbitofrontal - Right<br>Area of Rostralmiddlefrontal - Left<br>Area of Rostralmiddlefrontal - Right<br>Mean L1 in PosteriorCoronaRadiata on FA skeleton - Right<br>Mean RD in PosteriorCoronaRadiata on FA skeleton - Left<br>Mean L1 in PosteriorCoronaRadiata on FA skeleton - Left<br>Mean RD in GenuOfCorpusCallosum on FA skeleton<br>Mean ICVF inGenuOfCorpusCallosum on FA skeleton<br>Mean ICVF inRetrolenticularPartOfInternalCapsule on FA skeleton - Left<br>Mean RD in FornixCres_striaTerminalis on FA skeleton - Right |
| **Imaging** | O =1<br>Vascular problems diagnosed | **432 features with significant corrected correlation with UMAP embedding; Top 20 features:**<br>Area of Superiorfrontal - Left<br>Area of Superiorfrontal - Right<br>Area of Superiortemporal - Left<br>Mean RD in SuperiorCoronaRadiata on FA skeleton - Left<br>Area of Superiortemporal - Right<br>Mean RD in SuperiorFronto_occipitalFasciculus on FA skeleton - Left<br>Area of Lateralorbitofrontal - Left |
|  |  | Mean RD in SuperiorCoronaRadiata on FA skeleton - Right<br>Area of Lateralorbitofrontal - Right<br>Area of Rostralanteriorcingulate - Left<br>Mean RD in SuperiorFronto_occipitalFasciculus on FA skeleton - Right<br>Area of Precuneus - Right<br>Mean RD in PosteriorThalamicRadiation on FA skeleton - Right<br>Area of Middletemporal - Right<br>Mean RD in SuperiorLongitudinalFasciculus on FA skeleton - Right<br>Area of Medialorbitofrontal - Left<br>Mean RD in PosteriorThalamicRadiation on FA skeleton - Left<br>Mean RD in ExternalCapsule on FA skeleton - Right<br>Mean RD in SagittalStratum on FA skeleton - Right<br>Mean RD in SagittalStratum on FA skeleton - Left |
| **Environment** | / |  |
| **G-E** | O =13<br>• TRD<br>• Depression with anxious features<br>• Depression Neuroveg. Symptoms<br>• N depression episodes<br>• Health satisfaction<br>• Family relat. Satisfactions<br>• Friendship relat. Satisfactions<br>• Duration worst depression<br>• Frequency depressed days<br>• Impact on normal roles<br>• Thoughts of death<br>• Ever self harmed | **44 features with significant corrected correlation with UMAP embedding; Top 8 features:**<br>Alcohol intake frequency<br>Adulthood stress<br>Belittlement by partner or ex-partner as an adult<br>Someone to take to doctor when needed as a child<br>Felt hated by family member as a child<br>Felt loved as a child<br>Physical violence by partner or ex-partner as an adult<br>Physically abused by family as a child |
|  | Belief own life is Meaningful |  |
| **Genetics** | O =1<br>Diabetes diagnosed | **20 features with significant corrected correlation with UMAP embedding; Top 3 features:**<br>PRS ADHD<br>PRS AUTISM<br>PRS Anorexia Nervosa |

Abbreviations: G-E-Imaging, genetic-environment; G-E, genetic-environment; O, outcome; TRD, treatment-resistant depression; PRS, polygenetic risk scores; TRD, treatment resistant depression; FA, fractional anysotropy; RD, radial diffusivity; ICVF, Intracellular Volume Fraction; L1, L1 directions.

G-E-Imaging, E-Imaging, and Imaging predictors were predominantly driven by structural brain measures, including cortical areas in frontal/temporal regions and dMRI measures (particularly radial diffusivity (RD)) in corona radiata regions and Isotropic Compartment Volume Fraction (ICVF). G-E predictor was exclusively driven by environmental variables including alcohol intake, adult stress, childhood trauma, and social support factors. The G predictor was driven by PRS for ADHD, autism, and anorexia nervosa.

Community characterization using top-ranked features and socio-demographic factors revealed distinct phenotypic profiles aligning with observed health outcome risks (**Table S14**). For G-E features, COM1 and COM4 consistently emerged as highest-risk groups with distinct vulnerability profiles: COM4 showed high alcohol consumption patterns and moderate stress levels, and balanced demographics (mean age 56.46 years, 46.3% female), while COM1 (mean age 55.84, 65.5% female) was characterized by the most severe trauma profile including the highest adulthood stress, childhood physical abuse, and poorest childhood support. Within other feature sets, top-risk communities demonstrated distinct neurobiological patterns: G-E-Imaging communities at high risk for stress-related depression showed elevated RD (COM1, mean age 57.95 years, 89% female) or moderately reduced cortical areas (COM4, mean age 52.14, 93.6% female); E-Imaging communities with the highest vulnerability for cancer risk displayed reduced structural measures across regions (COM2, mean age 58.50, 72.1% female and COM1, mean age 56.73, 90.8% female); Imaging communities at high risk for vascular problems showed preserved-to-moderately reduced structural patterns (COM2, mean age 49.07, 90.3% female, with reduced cortical areas, COM4, mean age 50.58, 77% female, with elevated RD); G community at high risk for diabetes (COM3: mean age 54.06 years, 31.7% female, highest antidepressant usage amounting to 14.4%) showed elevated PRS for ADHD and autism. These patterns demonstrate that vulnerability profiles are both outcome-specific and modality-dependent, revealing multiple neurobiological pathways to risk. For most depressive symptoms, psychosocial, and self-harm outcomes, trauma-stress pathways (COM1-G-E) and substance-behavioral patterns (COM4-G-E) represent distinct risk mechanisms, alongside depressive stress-related and cardiometabolic outcomes showing neurobiological profiles associated with structural brain alterations (i.e., dMRI and sMRI) within the top-risk communities.

#### Replication datasets

For the GSRD replication set (**Table 6**), we extracted top-ranked features from the G-E predictor showing cross-cohort generalizability for self-harm behavior and depression subtypes with anxious features. The UMAP embedding patterns for G-E features are shown in **Figure S6**, supporting feature-level consistency in topological space construction across independent cohorts. Among 14 features showing significant correlation with UMAP embedding, working status emerged as the most relevant feature (p<0.004, Bonferroni corrected), partially replicating the UKB pattern where stress-related environmental variables dominated G-E rankings.

**Table 6.** Ranking of the most influential characteristics driving community structure from UKB-derived best predictors showing significant replication patterns across independent datasets of GSRD and HSR. The p-value significance threshold (Bonferroni corrected) associated with each feature is reported. Only feature sets that demonstrated significant community differentiation for their respective UKB-predicted outcomes are included.

| Predictor | Significant outcomes | Top-Ranked Features |
| --- | --- | --- |
| **GSRD** |  |  |
| **G-E** | Depression with anxious features<br>Ever self harmed | **14 features with significant corrected correlation with UMAP embedding; Top 1 features** ($p < 0.004$, Bonferroni corrected for 14 features):<br>Working Status |
| **HSR** |  |  |
| **Imaging** | Vascular problems | **232 features with significant corrected correlation with UMAP embedding; Top 13 features** ($p < 0.0002$, Bonferroni corrected for 232 features):<br>Mean thickness of Precuneus (Left)<br>Mean thickness of Insula (Left)<br>Mean thickness of Supramarginal (Left)<br>Mean thickness of Precuneus (Right)<br>Mean thickness of Insula (Right)<br>Mean thickness of Posteriorcingulate (Right)<br>Mean thickness of Lateralorbitofrontal (Left)<br>Mean thickness of Supramarginal (Right)<br>Mean thickness of Lateralorbitofrontal (Right)<br>Mean thickness of Inferiorparietal (Left)<br>Mean thickness of Superior temporal (Right)<br>Mean thickness of Superior temporal (Left)<br>Mean thickness of Parsopercularis (Right) |
| **G-E** | **TRD** | **16 features with significant corrected correlation with UMAP embedding; Top 2 features:**<br>Physically abused by family as a child (NS, $p=0.09$)<br>Felt hated by family member as a child ($p=0.002$, Bonferroni corrected for 16 features) |

Abbreviations: GSRD, The European Group for the Study of Resistant Depression (GSRD); HSR, Hospital San Raffaele ; G-E-Imaging, genetic-environment; G-E, genetic-environment; TRD, treatment resistant depression; NS, not significative.

For the HSR replication set, we focused on the two UKB-derived best predictors of TRD and vascular problems (**Table 6)**. The UMAP embedding patterns for these predictors are shown in **Figure S6**, demonstrating consistent topological space construction across cohorts. For the G-E predictor associated with TRD, feeling hated by family members emerged as the top significant feature (p<0.002, Bonferroni corrected), while childhood physical abuse by family showed trend-level significance. This replicated the environmental domain prominence observed in UKB, where trauma and family stress variables were highly predictive of TRD, while highlighting in HSR the greater effect of childhood family trauma. For the Imaging predictor associated with vascular problems, top features consisted predominantly of cortical thickness measures in precuneus, insula, supramarginal, posterior cingulate, and superior temporal regions (p<0.0002, Bonferroni corrected). Despite UKB feature ranking including both structural and dMRI measures, while HSR was dominated exclusively by CT measures, this result underlines a replication of structural brain patterns. Overall, these findings demonstrate feature-level consistency for UKB-derived best predictors showing cross-cohort replication, with environmental stress variables maintaining prominence in G-E predictors and neuroanatomical patterns showing stability across cohorts.

## Discussion

This work presents a novel data-driven application of TDA to cluster individuals with MDD based on diverse genetic, environmental, and brain characteristics that could anticipate mental and general health outcomes, including indices of disease severity, medical comorbidities, and TRD.

Our findings reveal clinically meaningful vulnerability pathways within MDD, with G-E combinations emerging optimal predictors for most mental health outcomes, driven predominantly by environmental stress-related variables that shape community structures identified. Distinct substance-behavioral and trauma-stress pathways were identified, representing risk trajectories that respectively predict anxious and neurovegetative symptoms versus TRD and episode severity.

TDA proved to be a robust and effective method for integrating multimodal data and for comparing their predictive capability, ultimately generating patient clusters (i.e., communities) showing clinical relevance in terms of health outcomes. Beyond its ability to integrate multimodal data, TDA offers the unique advantage of interpretability, since the Mapper output can be easily interpreted through graph theory, providing a network-based perspective on subject clusters and their underlying structure.

The identified clusters demonstrated selective but clinically meaningful replication across independent validation cohorts. Robust cross-cohort generalizability was confirmed specifically for self-harm behavior prediction, with consistent feature-level patterns observed and meaningful outcome-level statistical replication.

Our results highlight the utility of multimodal data integration as a superior approach for characterizing and stratifying individuals with MDD, supporting prior evidence on the multifactorial nature of heterogeneous psychiatric disorders like MDD (49). Multimodal data are increasingly adopted in mental health research to capture complex interrelationships across feature modalities and the heterogeneity inherent in psychiatric conditions (50). Recent studies have demonstrated the effectiveness of multimodal integration in predicting and characterizing various mental health disorders (19, 50–52), overcoming the limitations of unimodal approaches. As an unsupervised learning method, TDA shows promise for identifying meaningful subgroups within MDD characterized by homogeneous input features (genetic, environmental, and brain imaging) which may support the prediction of disease trajectories and treatment response.

### 3.1 Methodological innovation: graph-based community detection for MDD stratification

Our study introduces a novel methodological advancement in MDD research by implementing community-based prediction through TDA-derived graph structures, extending beyond the outcome-focused SAFE score analysis employed in our previous work (47). Traditional clustering approaches in psychiatry have employed diverse methodological strategies for MDD stratification, including k-means clustering applied to functional network connectivity and clinical symptoms (28), hierarchical clustering combined with canonical correlation analysis (CCA) for whole-brain functional patterns (53), and latent profile clustering analysis integrated with CCA and partial least squares for MDD symptom-based subtyping (54). Recent resting-state MRI studies have identified neurophysiological subtypes of MDD by applying clustering analysis together with dimensionality reduction via CCA or principal component analysis, integrating functional connectivity data, clinical ratings, and genetic information to enhance both clustering quality and interpretability of results (23, 55). Diverse approaches have been applied for MDD stratification extending beyond single-modal applications, including high-dimensional data clustering applied to clinical and genetic features for transdiagnostic psychiatric stratification including subtyping of MDD (56), multiple co-clustering models incorporating multimodal biomarkers including BOLD correlations, genetic markers, and clinical variables (23), and genetic distance-based hierarchical clustering using MDD-associated genetic markers (22). While these data-driven approaches have shown promise for identifying clinically meaningful MDD subgroups (5, 27, 57), systematic comparison of multimodal feature combinations and cross-cohort validation remain limited. Our community detection approach addresses these limitations by leveraging the natural graph structure produced by TDA Mapper, where nodes represent patient clusters and edges encode similarity relations, enabling the identification of densely interconnected patient communities with sparse connections to other subgroups (39, 58). This graph-theoretical framework offers distinct advantages over conventional clustering methods by capturing both local and global relationships in a unified approach, facilitating the integration of genetic, environmental, and neuroimaging data while maintaining interpretability through network-based visualization (59) and offering a richer view of patient stratification compared to classical clustering approaches. The effectiveness of graph-based community detection has been demonstrated across diverse biological contexts, including community organization comparisons across rs-fMRI and task-based networks (41), functional segregation analysis within resting-state fMRI networks (42), protein interaction network analysis for functional module identification (46) and differentiation of psychiatric patients from healthy populations (45). Notably, recent applications have specifically employed TDA Mapper followed by community detection algorithms: Birbiri et al. (2023) (43) used TDA Mapper combined with community detection on structural MRI and dMRI data to identify subgroups differing in cognitive and language skills, demonstrating how this approach can highlight features distinguishing communities and enable deeper understanding of subgroup-specific characteristics. Similarly, Schwertener et al. (2023) (44) applied a comprehensive TDA-community detection pipeline using rs-fMRI and clinical outcome data to identify topological groups, subsequently using these communities for classification tasks and clinical trajectory prediction.

This established methodological foundation underscores the interpretative and predictive potential of TDA-based graphs, particularly valuable in psychiatry where patient heterogeneity complicates diagnosis and treatment stratification. Our findings confirm the robust methodological foundation for translating TDA-derived communities into clinically actionable patient stratification tools, emphasizing the interpretative power of graph-theoretical methods.

### 3.2 Genes x Environment interactions

In UKB, G-E feature sets emerged as the most significant predictors across 13 clinical outcomes, including medical comorbidities associated with MDD, anxiety-related features, atypical neurovegetative symptoms, suicidal phenotypes, and TRD. Among these predictors, environmental features—particularly those related to stress and trauma—were especially prominent. Specifically, childhood traumatic experiences ranked among the most influential variables. Similarly, perceived stress in adulthood, frequency of alcohol consumption, and experiences of partner violence consistently appeared as top predictors across multiple outcomes.

Community characterization within the UKB dataset revealed distinct vulnerability profiles, with specific communities on G-E set consistently emerging as the highest-risk groups through different pathways. One community (COM4-G-E) demonstrated a substance-behavioral risk pattern characterized by elevated alcohol consumption alongside increased susceptibility to anxious and neurovegetative symptoms, while another (COM1-G-E) exhibited a more severe trauma-stress profile marked by higher rates of childhood physical abuse and compromised social-support networks, predicting treatment resistance and episode severity. This pattern suggests that different biological and psychosocial mechanisms contribute to MDD vulnerability, with alcohol-related behavioral patterns and trauma-stress pathways representing distinct risk trajectories within the MDD spectrum.

The importance of environmental stress-related determinants showed consistent replication across independent cohorts. In GSRD, working status emerged as the top-ranked feature within the G-E predictor that demonstrated robust cross-cohort replication for self-harm behavior. While UKB top-ranked features for the G-E predictor were dominated by childhood trauma and partner violence variables (felt hated by family members, physical abuse, belittlement by partner), in GSRD, working status emerged as the most relevant feature within the robustly replicated G-E predictor for self-harm behavior, representing another manifestation of psychosocial stress within the same environmental domain. Notably, this demonstrates a shift toward socioeconomic stress determinants while maintaining consistency within the same psychosocial vulnerability domain. It should be noted however that the specific stress/traumatic factors analysed in UKB were not available in GSRD, therefore we had to use a variable in a related domain. In HSR, the G-E predictor for TRD achieved statistical significance, and the feature ranking analysis successfully identified childhood family trauma (feeling hated by family members) as the predominant environmental determinant, directly replicating the trauma-stress pathway observed in UKB.

These findings demonstrate that psychosocial stressors maintain their central role in shaping outcomes in MDD across independent cohorts, with childhood trauma, interpersonal violence, and socioeconomic stressors all contributing to the environmental vulnerability pathway.

These results align with prior literature emphasizing the critical interaction between genetic predisposition and environmental stressors in etiopathogenesis of MDD. Environmental stressors seem to contribute to the onset of clinically relevant symptoms following the traditional Diathesis-Stress model (49, 60), suggesting that individuals with specific biological (e.g., genetic) and/or psychological (temperament) traits are more vulnerable to environmental influences (61). For instance, immune-related gene expression may mediate the relationship between emotional abuse and MDD (61) and polymorphisms in genes involved in cortisol signaling may interact with early-life stress to increase risk for depression and suicidality (62).

Building on these established findings, our results highlighted that G-E combinations achieved optimal predictive performance for most mental health outcomes, with environmental factors predominantly driving graph construction from the initial feature ranking phase. Genetic variables contributed minimally to topological structure, ranking low even during the UMAP dimensionality reduction step of TDA. This environmental predominance suggests that deepening understanding of environmental stressors may inform targeted early intervention strategies.

### 3.3 Brain features

Brain imaging features provided relevant findings for the creation of the clusters related to some of the investigated outcomes. Namely, G-E-Imaging was the best feature set for depression related to traumatic or stressful events; E-Imaging for the outcome of lifetime cancer diagnosis; and finally, imaging alone resulted the best predictive set for vascular disease in MDD. For all three outcomes, frontotemporal regions and measures of RD of the dMRI appeared as the most important features.

Community characterization within the UKB dataset using feature sets including brain imaging revealed neurobiological vulnerability patterns across different clinical outcomes. High-risk for stress-related depression (COM1- and COM4- G-E-Imaging) showed preserved RD in WM tracts alongside intermediate-to-reduced cortical values. Similarly, communities associated with cardiometabolic outcomes (i.e., vascular problems and cancer diagnoses, COM2-, COM1-E-Imaging and COM2-, COM4-Imaging) demonstrated comparable neurobiological patterns, showing intermediate-reduced cortical areas while maintaining white matter structural integrity.

This suggests that both stress-related depression and cardiometabolic vulnerability pathways may involve similar compensatory mechanisms that preserve white matter microstructural organization.

Partial replication of anatomical patterns within the HSR independent cohort was observed, particularly for vascular comorbidities outcome. The imaging predictor achieved statistical significance, and feature-level analysis revealed consistent neuroanatomical patterns. Specifically, cortical thickness measures in precuneus, insula, and superior temporal regions successfully replicated the structural patterns identified in UKB. This replication confirms the stability and cross-cohort generalizability of neuroanatomical markers for vascular comorbidities in MDD.

The frontotemporal alterations identified in our analysis may reflect shared neurobiological pathways underlying both MDD and vascular comorbidities, rather than MDD-specific mechanisms. Emerging evidence suggests that depression and cardiovascular disease may represent distinct clinical manifestations linked by common central autonomic network (CAN) dysfunction (63, 64). The CAN, comprising frontal, cingulate, and insular cortices alongside brainstem structures, serves as the primary neural substrate for cardiovascular control and has been implicated in both depressive symptomatology and cardiovascular regulation (65, 66). Recent studies demonstrate that acute inflammatory challenge compromises CAN function, creating neurobiological vulnerability that manifests as both autonomic dysregulation and increased cardiovascular risk (64). The fronto-limbic neural loop contributes to depression-related disturbances in autonomic regulation and neuroendocrine responses via connections with visceral control structures including the hypothalamus and brainstem (63). This pathophysiological overlap is mediated through CAN dysfunction, acute inflammatory responses, and altered brain-heart axis communication that simultaneously affect cardiac function and mood regulation (63, 64). The frontotemporal cortical alterations observed in our high-risk communities may therefore represent a central endophenotype of cardiovascular dysfunction that predisposes to both vascular comorbidities and depressive symptoms through disrupted brain-heart axis communication (63). This conceptual framework suggests that structural brain changes in MDD with vascular comorbidities reflect integrated cardio-cerebral pathology rather than independent disease processes, supporting the clinical utility of neuroimaging markers for identifying individuals at risk for both psychiatric and cardiovascular outcomes.

### 3.4 Study limitations and future perspectives

TDA application requires the tuning of multiple parameters, including resolution, gain, and overlap settings, which can influence the resulting graph topology and community structure (34, 35). Although we adopted well-established approaches for TDA parameter configuration in line with previous research (30, 47), we did not perform a systematic quantitative evaluation for parameter selection. Rather, we employed fixed settings (R=15, G=80% inter-bin overlap) derived from our prior investigations (47), applied to the same domains of genetic, environmental, neuroimaging sets, and validated TDA protocols that have shown efficacy in neuropsychiatric research contexts. While this represents a widely accepted methodology, the approach depends on parameter choices informed by previous experience rather than data-driven optimization specific to each dataset. Although extensive parameter space exploration using performance metrics could be conducted (67), hyperparameter selection would still involve subjective decisions and context-specific adaptations, which may limit cross-study generalizability. Future research should investigate the balance between automated parameter optimization and consistent methodological approaches (67), establishing systematic selection procedures that maintain both analytical rigor and reproducibility across diverse samples.

Secondly, we implemented a systematic approach to community detection by quantitatively comparing multiple state-of-the-art algorithms, including Louvain modularity optimization, Label Propagation, and Fastgreedy approaches, with algorithm selection based on comprehensive evaluation of coverage, modularity, and overall network performance metrics (39, 40). While Louvain demonstrated best performance across most of our feature sets, we acknowledge that the choice of community detection algorithm can influence results, and testing additional methods (e.g., Leiden, Infomap, or Spectral clustering) might have revealed superior algorithms that could outperform Louvain for specific datasets. Although we selected algorithms based on rigorous performance evaluation (40, 46, 68), and these methods have demonstrated success in network analysis applications, there is no universal guarantee that the resulting communities provide the optimal functional description of the underlying system. The field lacks consensus on universally optimal community detection approaches, as performance depends on the specific network properties and topological characteristics of each dataset. Future investigations should explore additional community detection algorithms to better understand how methodological choices influence community identification across different data structures and network configurations.

In addition, considering that our community-detection algorithms comparison falls within Intrinsic Quality Measures (IQM) evaluation approaches (69), we need to evaluate the quality of clusters without reference to external information, thus relying on properties as modularity. Although we consider properties defined as important by the literature studies on network structures (i.e., modularity, coverage, performance), future studies should implement a more comprehensive evaluation strategy incorporating additional network parameters or adopting approaches that focus exclusively on modularity optimization for community detection.

Furthermore, to ensure robust community identification suitable for subsequent statistical testing, we implemented an arbitrary minimum threshold of 5 core samples per community. This criterion was designed to maintain adequate statistical power for meaningful between-group comparisons and reliable community characterization. However, this decision may systematically exclude potentially relevant smaller subgroups with distinct biological or clinical characteristics.

Our novel two-stage feature selection strategy combines UMAP-guided dimensionality reduction with community-based validation to identify the most relevant features for community differentiation. While this approach showed consistent results across datasets, different embedding techniques (e.g., t-SNE, PCA) might produce alternative low-dimensional representations, potentially affecting both TDA graph construction and the subsequent feature ranking pipeline, leading to different biological interpretations.

Despite systematic efforts to harmonize features and outcome measures across datasets, inherent differences in data collection protocols, assessment instruments, and variable definitions between UKB and replication cohorts may have introduced systematic variation affecting community-outcome comparability.

Environmental and clinical measures were assessed using different questionnaire versions and temporal frameworks across sites, potentially influencing the detection and interpretation of community-based associations.

Neuroimaging validation presented additional challenges despite implementing standardized processing pipelines derived from established UKB protocols. GSRD lacked neuroimaging data entirely, restricting imaging-based community validation to HSR alone. The HSR dataset lacked resting-state and task-based functional MRI modalities due to acquisition protocol differences and sample size constraints, preventing comprehensive validation of multimodal neuroimaging findings. Cross-site differences in scanner hardware, acquisition parameters, and imaging protocols across centers, while addressed using the same software for pre-processing approaches, may have introduced technical variance affecting the reproducibility of neuroimaging-derived community structures. These methodological constraints limited the comprehensive validation of the neuroimaging components of our multimodal framework. Despite these limitations, the replication cohorts demonstrated reasonable demographic and clinical homogeneity with the UKB sample.

To conclude, the MDD individuals considered in this study may not be representative of the general MDD population. UKB is enriched in females, elderly, wealthier, and more educated individuals with respect to the general UK population (70) and represents a general population longitudinal cohort rather than clinical cross-sectional samples. This sampling difference may limit our ability to characterize certain outcomes, particularly medical comorbidities and treatment resistance, as our MDD sample likely represents a milder subpopulation within UKB compared to severe clinical depression cases.

## Materials and Methods

We analyzed three MDD cohorts: UKB, the European Group for the Study of treatment Resistant Depression (GSRD, N=1,017) (71), and a cohort from Hospital San Raffaele (HSR, N=87) (72). All participants met MDD diagnostic criteria with detailed inclusion/exclusion criteria and a general description of the samples reported in the Supplementary Methods.

In each cohort, we extracted participants with the available features of interest, i.e., G-E and G-E-I. In the UKB, the G-E cohort included N=20,715 individuals with complete genetic and environmental data, while the G-E-I cohort included N=3,044 participants. The GSRD cohort did not include brain imaging but G-E features (N=1,017), while the HSR cohort included N=87 individuals in the G-E subset and N=71 in the G-E-I subset. The final feature dimensions were obtained by removing features with excessive missing data, then selecting subjects with complete data for remaining features within each set. The included subjects were selected according to any available measure/assessment.

### 4.1 Measures

#### UKB predictive features

From the G-E cohort, multiple feature sets were subsequently extracted, including gene-only (G, P=20 features), environment-only (E, P=33 features), and gene-environment combination (G-E, P=53 features). From the G-E-Imaging main cohort, imaging-related feature sets were derived, including imaging-only (P=606 features), environment-imaging (E-Imaging, P=639 features), and gene-environment-imaging combinations (G-E-Imaging, P=659 features). The list of variables included in each feature set of UKB sample is reported in the corresponding column of **Table 3**.

Brain imaging features included (i) sMRI measures comprising regional subcortical and cortical volumes, cortical thickness, and cortical surface areas; (ii) dMRI regional measures including fractional anisotropy, diffusivity indices, and microstructural measures; (iii) rs-fMRI measures of functional connectivity strength of networks of interest (see **Table S1**); and t-fMRI activation measures from emotional processing paradigms. Details on the MRI data pre-processing and feature extraction are reported in the Supplementary Methods. Genetic features consisted of PRSs for psychiatric disorders, behavioral traits, and immune-cardiometabolic conditions estimated using PRS-CS-auto (73) and calculated using the score function in PLINK 2.0 (74) in participants of European ancestry. Each PRS was adjusted for ancestry-relevant population principal components, genotyping batch, and center of recruitment before inclusion in the analyses (see **Table S2** for a description of the GWAS summary statistics). Environmental features encompassed both lifetime experiences and variables partly depending on them, including personality traits, social support, substance use patterns, lifestyle factors, and traumatic events (47). A detailed description of the variables included in environmental set is reported in **Table S3**. Confounding variables included age, sex, ethnicity, and antidepressant medication status (yes/no). Detailed descriptions of socio-demographic variables can be found in **Tables S4 and S5**.

#### UKB health-related outcomes

As reported in **Table 2**, we considered 18 health outcomes comprising variables related to depression severity, including TRD, presence of atypical neurovegetative symptoms or anxious features, depression related to stressful or traumatic events, number of depressive episodes, duration of worst depression, frequency of depressed days, and impact on normal roles. Self-harm and suicidal thoughts or behaviors were assessed through thoughts of death and self-harm history. Psychosocial functioning was evaluated via health satisfaction, family relationship satisfaction, friendship relationship satisfaction, work/job satisfaction, and belief that one’s own life is meaningful. Finally, we considered somatic comorbidities including cancer, diabetes, and vascular problems diagnosis. A detailed description of each outcome variable is reported in **Table S7**.

#### Replication cohorts

Predictive features and health-related outcomes as consistent as possible with the UKB sample were extracted. Features and health-related outcomes harmonization across UKB and replication cohorts was conducted to ensure comparability, with detailed matching between replication sets and UKB variables presented in **Tables S8-S9**. These included the available PRSs, environmental factors measured using partly different questionnaires but reflecting the same domains(e.g., trauma exposure, lifestyle factors), the same neuroimaging features from sMRI and dMRI only for HSR, and comparable health-related outcomes The predictive variables included in each feature set of the replication cohorts are reported in **Table 3** in the corresponding column (i.e., GSRD or HSR), while **Table 2** reports the health-related outcomes.

In the GSRD sample, feature sets included environmental variables (E, P=5) covering marital status, occupation, smoking habits, social support, and abuse experiences; PRSs (G, P=18); and gene-environment features (G-E, P=23). Age and sex (P=2) were concatenated to each predictive feature set. Health-related outcomes comprised 6 measures coherent with UKB: TRD, depression with anxious features, thoughts of death during the current depressive episode, suicidal and self-harm behaviors, somatic disorders, and duration of the current depressive episode.

In the HSR sample, for whom imaging data was available, feature sets included environmental variables (E, P=12), PRSs (G, P=19), gene-environment variables (G-E, P=31), neuroimaging features from sMRI and dMRI (Imaging, P=344) and concatenation of all features (G-E-Imaging, P=375). Age, sex, ethnicity, and antidepressant medication status were concatenated to each predictive feature set. Health-related outcomes comprised 11 measures coherent with UKB: TRD, depression with anxious and atypical neurovegetative features, number of depressive episodes, thoughts of death during the current episode, suicidal and self-harm behaviors, duration of current depressive episode sense of personal meaningful life, and somatic comorbidities (cancer, diabetes, vascular problems). For replication samples, a detailed description of the predictive features is reported in **Table S6** and **S8** while a detailed description of health-related outcome variables is reported in **Table S9**.

### 4.2 Data analysis

#### TDA Mapper application and Community-detection

For UKB, different TDA Mapper graphs were constructed by separately employing the available gene, environment, and imaging feature sets, alone and in combination: (i) G, (ii) E, (iii) G-E, (iv) Imaging, (v) E-Imaging, and (vi) G-E-Imaging). TDA Mapper graphs, extracted using the *kepler library* (*75*), were clustered in communities employing community-detection algorithms. Confounding variables were concatenated to all feature sets to compare the the predictive capability of the latter while controlling for any interactions with these confounding effects. The pipelines of TDA graphs construction and community detection described as follows were applied coherently to UKB and replication sets. For the replication sets, TDA graphs were built considering the feature sets that were marked as best predictors for the selected outcomes, based on their availability. The HSR replication set included all feature sets of the UKB set, the GSRD replication set included only (i) G, (ii) E, (iii) G-E feature sets.

#### TDA Mapper graph construction

For each feature set, a TDA graph was constructed using the Mapper algorithm. The feature space was established by defining each selected feature set as the subjects’ space S, employing Euclidean distance as the primary similarity measure between data points, consistent with established methodologies for multimodal feature integration in TDA (30, 34, 37).

Prior to the analysis, all features were preprocessed through robust scaling utilizing median and interquartile range statistics to minimize outlier effects and harmonize diverse feature modalities (environmental, genetic, and imaging variables) in distance calculations. This preprocessing strategy maintains the integrity of underlying data relationships and preserves subject-specific patterns more effectively than conventional normalization techniques when working with heterogeneous biological measurements, aligned with established TDA methodological frameworks (76).

The Mapper algorithm was implemented with Uniform Manifold Approximation and Projection for dimensionality reduction (UMAP) configured for 2-dimensional embedding to reduce complexity while maintaining topological characteristics. UMAP was selected for its capacity to preserve local neighborhood relationships and global topological structure, critical for multimodal data integration (77, 78). Following projection, the mean and variance of the transformed data matrix were calculated. By considering a 2-dimensional UMAP filtering, the actual covering procedure generates overlapping rectangular regions, facilitating data compression and noise reduction (47).

The resulting data were partitioned into overlapping intervals using cover parameters: Resolution (R=15, number of bins) and Gain (G=80%, inter-bin overlap), defined according to a previous related work (47). These parameters selection was conducted through systematic evaluation of R-G combinations using a G-E-Imaging feature set, with selections based on topological consistency (69).

Clustering was applied to cluster subjects within covering bins, tailoring the method to the dataset characteristics: Density-based spatial clustering (DBSCAN) was implemented for smaller UKB feature sets (G-E-Imaging, E-Imaging, Imaging) and all replication feature sets, due to enhanced performance with variable density distributions. Hierarchical density-based spatial clustering (HDBSCAN) was deployed only for larger UKB feature sets (G, E, G-E) for superior handling of high-dimensional spaces. Both approaches were chosen for their adaptive density handling and parameter-free cluster identification (79, 80). The final graph topology was established by connecting overlapping clusters through edge placement between bins containing shared subjects.

#### Community detection within the TDA graph

Following TDA graph construction, community detection algorithms were systematically applied to partition the TDA graphs into meaningful and robust communities. Multiple state-of-the-art algorithms were tested and compared, including Louvain modularity optimization, Label Propagation, Fastgreedy approaches, with best algorithm selection based on comprehensive evaluation of key quality metrics including modularity, coverage, and performance as implemented in *NetworkX library* (81–83). This comparative approach ensured that identified communities were both statistically robust and biologically meaningful in representing the underlying data structure and subgroups. Communities containing fewer than 5 samples were considered statistically unreliable and systematically excluded from subsequent analyses. Following community identification, core samples— i.e., individuals assigned to a single community with 100% membership confidence— were extracted and utilized exclusively for all downstream analyses, as these represented the most reliable community assignments while excluding samples located on community boundaries or in overlapping zones between multiple communities.

### 4.3 Community-based stratification of outcomes

In the UKB sample, we tested whether the identified communities achieved effective stratification in terms of the health-related outcomes to evaluate community clinical relevance. Significant differences in outcomes across communities were tested for each feature set using Kruskal-Wallis (KW) tests for ordinal/continuous variables and Chi-squared tests (Chi2) for dichotomous variables. Multiple comparisons correction was applied using Bonferroni adjustment (p<0.05, corrected for each outcome across the number of feature sets tested, N=6). Parallelly, effect sizes were calculated using Eta² for variables tested with KW and Cramer’s V for variables tested with Chi2 tests (84–86). The statistics obtained rated the community-based predictive capability of the TDA graph in terms of each of the health-related outcomes.

### 4.4 Comparison of predictive capabilities of feature sets

In the UKB dataset, we identified which feature set was the best predictor for each outcome. The best predictor was identified as the feature set showing the greatest statistical significance across communities.

Following the identification of the best predictor for each outcome, in the selected TDA graph, community-based outcome risk ranking was performed by extracting the percentage of subjects at "high risk" within each community. High-risk categories were defined: (i) for binary outcomes, as the presence of the risk condition; (ii) for ordinal scales, as the set of categories reflecting negative outcomes (further details can be found in **Table S12**). This approach enabled identification of high-risk communities and lower-risk communities, providing meaningful stratification of MDD in terms of clinical outcomes.

### 4.5 Best predictors: test of replicability

For independent test sets (GSRD and HSR), we specifically evaluated whether the best predictor feature set identified in UKB for each outcome was able to differentiate outcome subgroups also in these independent samples. To this end, the community-based stratification of outcomes described in section 4.3 was employed.

### 4.6 Best predictors: feature ranking pipeline

For both UKB and the replication sets, a feature ranking pipeline was developed and applied to identify the most influential variables driving TDA graph construction and community differentiation. For each outcome separately, in the corresponding best predicting feature set (e.g., G-E), we implemented a novel two-stage feature ranking approach. We extracted features defining the TDA space and then filtered them to retain only those that mostly differentiated TDA graph communities.

#### Step 1: UMAP-guided feature selection

This step identified the features most responsible for defining the low-dimensional space that forms the foundation for TDA graph topology and community structure. The first 2 UMAP components (UMAP1 and UMAP2) were extracted from the high-dimensional dataset, and linear correlations were computed between them and each feature, yielding correlation coefficients and p-values. Multiple testing correction using False Discovery Rate (FDR) with Benjamini-Hochberg procedure was applied separately to correlations with UMAP1 and UMAP2 components (p<0.05, FDR-corrected). Top features were selected based on significant correlation with both UMAP components: only features showing significant correlations with both UMAP1 and UMAP2 were retained; then, from these significant features, the top-rank features were selected based on correlation strength, specifically, for each UMAP component, the top 10% most correlated features for datasets with ≤100 features, or the top 10 features for larger datasets.

#### Step 2: Community-based feature selection

Features selected in Step 1 underwent community-based validation using KW tests or Chi2 tests to assess significant differences across TDA-derived communities (p<0.05, Bonferroni corrected for the number of features selected in Step 1). Features with significant omnibus test results were retained as the final discriminative feature set. This second step of the pipeline captures the global and local TDA structure by prioritizing features that not only drive the overall UMAP embedding but also reflects the community differentiation, ensuring the consideration of only those features that strongly explain both graph and community structures.

In both UKB and replication datasets, these steps were applied to the best predictive feature sets for the health-related outcomes. In replication datasets (GSRD and HSR), we validated the top-ranked features from UKB’s best predictor sets for each outcome, providing a focused assessment of feature ranking consistency and clinical relevance across independent populations.

### 4.7 Best predictors: community characterization and risk-ranking

For the best predictive feature set obtained for each outcome, after performing community-based outcome risk ranking by determining the community with the highest risk for each outcome (described in 4.4), we characterized each UKB community across additional key dimensions: (i) top-ranked features obtained after Step 2 of the feature ranking pipeline, and (ii) socio-demographic profiles including age, sex, ethnicity, and antidepressant medication status. This multi-dimensional characterization approach enabled a systematic characterization of community profiles in terms of input features, clinical outcomes and confounders.

## Supporting information

Supplementary Materials related to the main manuscript.

## Data Availability

There are restrictions prohibiting the provision of data in this
manuscript. The data in this manuscript were obtained from a third party, UK Biobank,
upon application. The genetic, environmental, brain imaging and clinical outcomes data
have been deposited with the UK Biobank and are freely available to approved researchers, as has been done with other datasets to date. Interested parties can apply for data from UK Biobank directly, at http://www.ukbiobank.ac.uk. UK Biobank will consider data applications from bona fide researchers for health-related research that is in the public interest. By accessing data from UK Biobank, readers will be obtaining it in the same manner as we did. The central code of the main work is available upon request at https://osf.io/cphj6/?view_only=cc5f359232494a828ad4807f544ae1eb.

## Acknowledgments

This research has been conducted using the UK Biobank Resource under Application Number 56514. “Stratification of health outcomes in mood disorders”. The study was supported by the Italian Ministry of Health (DEPTYPE project, grant n. GR-2019-12370616). PB was partially supported by grants from the Italian Ministry of Education and Research - MUR (‘Dipartimenti di Eccellenza’ Programme 2023–27 - Dept. of Pathophysiology and Transplantation, Università degli Studi di Milano), the Italian Ministry of Health (Hub Life Science-Diagnostica Avanzata, HLS-DA, PNC-E3-2022-23683266– CUP: C43C22001630001 / MI-0117; Ricerca Corrente 2025), and by the ERANET Neuron JTC 2023 (ERP-2023-23684211 - ResilNet) and Eranet Neuron JTC 2024 (ER-2024-23684536 - BRAWO Project). EM was partly supported by the Italian Ministry of University and Research (PRIN 2022 PNRR, grant n. P20229MFRC). The European Group for the Study of Resistant Depression (GSRD) obtained an unrestricted grant sponsored by Lundbeck A/S. We acknowledge the contributions of the GSRD network members: Siegfried Kasper, Joseph Zohar, Daniel Souery, Stuart Montgomery, Panagiotis Ferentinos, Dan Rujescu, and Julien Mendlewicz. The sponsor played no role in designing the study, data collection and analyses, interpretation of the data, writing of the manuscript, and in the decision to submit the research for publication.

## Notes

### Competing Interest Statement

The authors have declared no competing interest.

### Author Declarations

All procedures were performed in compliance with relevant laws and institutional guidelines. Informed consent was obtained for studies on human subjects. The institutional committee of IRCCS Ospedale San Raffaele has approved the HRS study (date 16/12/2020; ref. number, 211/INT/2020). The European Group for the Study of Resistant Depression (GSRD) study protocol was approved by the Ethics Committee of the coordinating center at Hopital Erasme, Cliniques universitaires de Bruxelles (Universite Libre de Bruxelles, approval number B406201213479), Belgium, and the local ethical committees of all participating centres. The UKB obtained ethics approval from the North West Multi-centre Research Ethics Committee with approval number 11/NW/0382.

