## Supplementary Materials related to the main manuscript. for "Topological data analysis communities reveal gene-environment-brain subtypes of major depression in UK Biobank and multi-site cohorts"

**This PDF file includes:**

Supplementary information text: Methods	p. 2-4

Fig. S1 to S6	p. 5-8

Tab. S1 to S14	p. 9-30

Supplementary references.

Supplementary Information Text

**Methods.**

**Participant Characteristics and Diagnostic Criteria.**

***UKB cohort***: The UKB (UKB) is a population-based cohort from the United Kingdom including approximately 500,000 individuals, with comprehensive collection of biomarkers as longitudinal environmental, lifestyle, activity, genetic, multimodal neuroimaging data, alongside with health-related information.

We included participants with MDD diagnosis with any available measure/assessment, in accordance to the following MDD diagnostic criteria: In detail, we considered: 1) diagnosis of a depressive disorder according to primary care records, considering diagnostic codes previously described (at least one diagnostic code for depression, n=37,394) (Fabbri et al., 2021); 2) MDD defined by the Composite International Diagnostic Interview Short Form (CIDI-SF) (n=34,870), which was part of the Mental Health Questionnaire (MHQ) (Fry et al., 2017); 3) hospital diagnoses (ICD-10 codes F32-F33) (n= 17,271); 4) Smith et al. definition (n=30,876 (Davis et al., 2020) . Exclusion criteria encompassed participants with bipolar, psychotic, or substance use disorder diagnoses identified through primary care records, hospital ICD-10 codes, and/or MHQ data (Fabbri et al., 2021).

**European Group for the Study of treatment Resistant Depression (GSRD) cohort: The GSRD cohort** (Kautzky et al., 2025) comprised 1148 patients with MDD recruited through a multicenter, cross-sectional study including hospitalized and outpatient participants. **Diagnostic criteria** included MDD diagnosis according to DSM-IV-TR criteria confirmed using the Mini International Neuropsychiatric Interview (MINI). **Inclusion criteria** encompassed: (a) receipt of at least one antidepressant during current MDD episode (≥4 weeks at adequate dose); (b) Montgomery-Åsberg Depression Rating Scale (MADRS) score >22 at current episode onset. **Exclusion criteria** included any primary psychiatric disorder other than MDD, substance use disorders (except nicotine and caffeine) in the preceding 6 months, and any condition interfering with informed consent or study assessments.

**Hospital San Raffaele (HSR) cohort: The HSR cohort (Colombo et al., 2024)** recruited 157 participants admitted to the inpatient unit. Patients were first screened to have a score higher than 8 at the 21-Hamilton Depression Rating Scale (HDRS-21) (Hamilton, 1960), then evaluated according to the fifth edition of the Diagnostic and Statistical Manual of Mental Disorders (DSM-5) criteria, assessed using the Structured Clinical Interview to evaluate the presence an ongoing depressive episode. All the participants had to be between 18 and 65 years old. **Exclusion criteria** comprised comorbidity with other psychiatric pathologies, first-onset depressive episode, pregnancy, major medical and neurological disorders, and substance abuse in the last 6 months. These final sample considered for the analysis, together with the features considered were obtained through a two-step missing data handling strategy: first, features with excessive missing data were systematically removed across the sample; secondly, subjects with complete data for the remaining features within each set were selected obtaining **N=87 individuals in the Genetic-Environment (G-E) subset and N=71 in the Genetic-Environment-Imaging (G-E-Imaging) subset.**

**Pre-processing of brain imaging data.**

***UKB cohort:*** The processing and quality control (QC) pipelines applied to brain imaging data were previously described (Alfaro-Almagro et al., 2018). Briefly, part of the magnetic resonance imaging (MRI)-derived features extraction’s pipelines and related parameters were reported as follow for UKB.

***sMRI****.* T1-weighted imaging was performed by using a three-dimensional magnetization-prepared rapid acquisition with gradient echo (MPRAGE) sequence (resolution: 1.0 x 1.0 x 1.0 mm; field-of-view matrix: 208 x 256 x 256; inversion time, 880 msec; repetition time, 2000 msec). T1-weighted data were first segmented by using FAST (version 4.1; Automated Segmentation Tool in FMRIB) to extract gray matter, white matter, and cerebrospinal fluid. Subcortical structures were extracted by using FIRST (version 5.0; Integrated Registration and Segmentation Tool in FMRIB). Cortical regional volumes, cortical thickness and surface area were bobtained using Freesurfer according on Desikan-Killiany atlas (UKB Data fields within the Category: 1101, [Regional grey matter volumes (FAST)](https://biobank.ndph.ox.ac.uk/showcase/label.cgi?tk=yZjRoJFA3dq4WUv7alKluuMBHS4bpDM7189922&id=1101); 1102, [Subcortical volumes (FIRST)](https://biobank.ndph.ox.ac.uk/showcase/label.cgi?tk=yZjRoJFA3dq4WUv7alKluuMBHS4bpDM7189922&id=1102); 196, [Freesurfer DKT](https://biobank.ndph.ox.ac.uk/showcase/label.cgi?id=196)).

***dMRI****.* Diffusion weighted MRI (dMRI) was performed using echo-planar, single-shot Stejskal-Tanner pulse sequence (echo time msec, 92) to obtain 36 sections (resolution, 2.0 x 2.0 x 2.0 mm; field-of-view matrix: 104 x 104 x 72 matrix) in 50 distinct diffusion-weighted directions (*b* values, 1000 and 2000 sec/ mm2). dMRI in individual white matter tracts were extracted using FMRIB Software Library (FSL) tools (<http://www.fmrib.ox.ac.uk/fsl>). dMRI variables included fractional anisotropy (FA), orientation dispersion index (ODI), measures such as Intracellular Volume Fraction (ICVF), the Isotropic Compartment Volume Fraction (ISOVF), radial diffusivity (RD) and L1 directions. All dMRI measures were averaged within 48 distinct tract ROIs defined using the Johns Hopkins University tract atlas, with measures calculated separately for left and right hemispheres. (UKB Data fields within the Category 134, [dMRI skeleton](https://biobank.ndph.ox.ac.uk/showcase/label.cgi?tk=yZjRoJFA3dq4WUv7alKluuMBHS4bpDM7189922&id=134)).

***rs-fMRI***. rs-fMRI was performed using gradient echo echo-planar imaging (GE EPI) (resolution: 2.4 x 2.4 x 2.4 mm; field-of-view matrix: 88 x 88 x 64; duration time, 6 min; repetition time, 0.735 sec; echo time, 39 msec; flip angle, 52°). rs-fMRI network matrices were employed, extracted from the imaging-derived phenotypes that were processed by the UKB imaging project team. Data preprocessing, group independent component analysis (ICA) parcellation, and connectivity estimation were carried out using FSL packages (http://biobank.ctsu.ox.ac.uk/ crystal/refer.cgi?id=1977) according to the UKB pipelines. Network matrices obtained using group-ICA with dimensionality of D=100 independent component (ICs) and non-artificial components were considered. This identified 55 components in 100-D that remained for further analysis. Connections between pairs of ICs were extracted by considering partial correlation as measure of functional connections among components obtaining a 55x55 partial correlation matrix for each individual. From the matrix of connectivity estimates extracted at subject-level with dimension 55 ICs x 55 ICs were extracted the parameters of strength of positive (Spos) and negative weights (Sneg) for each network “node”, resulting in 55 Spos and 55 Sneg measures. Hence, each of the 55 ICs were visualized based on the UKB’s online visualization tool ([https://www.fmrib.ox.ac.uk/datasets/UKB/group_means/rfMRI_ICA_d100.html](https://www.fmrib.ox.ac.uk/datasets/ukbiobank/group_means/rfMRI_ICA_d100.html)) and grouped into 9 typical large functional networks, in accordance with previous studies (Jiange et al., 2023) (UKB Data fields within the Category 203, [Resting functional brain MRI](https://biobank.ndph.ox.ac.uk/showcase/label.cgi?tk=yZjRoJFA3dq4WUv7alKluuMBHS4bpDM7189922&id=111)).

***t-fMRI***. t-fMRI was performed using GE EPI (resolution: 2.4 x 2.4 x 2.4 mm; field-of-view matrix: 88 x 88 x 64; duration time, 4 min; repetition time, 0.735 sec; echo time, 39 msec; flip angle, 52°). Task-based activation during implicit emotion processing of the Hariri faces/shapes “emotion” task (Hariri et al., 2002) were extracted. Specifically, the participants were presented with blocks of trials and asked to decide either which of two faces presented on the bottom of the screen match the face at the top of the screen, or which of two shapes presented at the bottom of the screen match the shape at the top of the screen. The faces have either angry or fearful expressions. These variables include Median blood-oxygen-level-dependent (BOLD) effect and 90^th^ percentile (in group-defined amygdala activation mask) for faces-shapes contrast, Median BOLD effect and 90^th^ percentile (in group-defined mask) for faces activation, Median BOLD effect and 90^th^ percentile (in group-defined mask) for faces-shapes contrast, Median BOLD effect and 90^th^ percentile (in group-defined mask) for shapes activation (UKB Data fields within the Category 106, [Task functional brain MRI](https://biobank.ndph.ox.ac.uk/showcase/label.cgi?tk=yZjRoJFA3dq4WUv7alKluuMBHS4bpDM7189922&id=106)).

***HSR cohort:*** HSR data for neuroimaging were processed according to UKB protocols, adapting them according to the specific acquisition parameters of HSR data. Below we reported MRI acquisition parameters.

A sample of 59 patients was acquired with Gyroscan Intera, Philips, Netherlands employing a 8 channels. For sMRI, SENSE head coil (T1-weighted MPRAGE sequences: TR 25.00 ms, TE 4.6 ms, field of view FOV=230 mm,91 matrix=256×256, in-plane resolution 0.9×0.9 mm, yielding 220 transversal slices with a thickness of 0.8 mm) (sMRI). For dMRI, SE EPI sequences (TR/TE=9000/58 ms, FoV (mm) 232(ap), 126 (fh), 240.00 (rl); acquisition matrix = 112×85; voxel acquisition 2.14×2.73×2.3; 55 contiguous, with in-plane voxel size 1.88×1.88 mm; SENSE acceleration factor=2; 1 b0 and 35 non-collinear directions of the diffusion gradients; b value=900 s/mm2) were used.

A sample of 70 patients was instead acquired with an Ingenia CX, Philips, The Netherlands using a 32-channel sensitivity encoding. For sMRI, SENSE head coil (T1-weighted MPRAGE sequence: TR 8.00 ms, TE 3.7 ms, field of view FOV = 256 mm, matrix = 256 x 256, in-plane resolution 1 x 1 mm, yielding 182 transversal slices with a thickness of 1 mm). For dMRI, SE EPI sequences (EPI factor= 43; TR/TE=5900/78 ms, FoV (mm) 232 (ap), 129 (fh), 240.00 (rl); acquisition matrix 112×85; 56 contiguous, 2.3-mm thick axial slices reconstructed with in-plane pixel size 1.88×1.88 mm; SENSE acceleration factor= 2; Multiband acceleration factor= 2; ten b0 and 96 non-collinear directions of the diffusion gradients: 60 b values=2855 s/mm2, 6 b values=700 s/mm2, 30 b values=1000 s/mm2) were acquired. Fat saturation was performed to avoid chemical shift artefacts.

**Genotyping, quality control and imputation.**

**UKB cohort:** Genome-wide genotyping was performed all UKB was performed using two highly overlapping arrays covering ~600.000 markers. Autosomal genotype data underwent centralized quality control to adjust for possible array effects, batch effects, plate effects, and departures from Hardy-Weinberg equilibrium (HWE) (Bycroft et al., 2018). SNPs were further excluded based on missingness (> 0.05) and on Hardy Weinberg equilibrium (p <10-8). Individuals were removed for high levels of missingness (> 0.05) or abnormal heterozygosity (as defined during centralized quality control), relatedness of up to third-degree kinship (Manichaikul et al., 2010) (KING r < 0.044) or phenotypic and genotypic gender discordance. Population structure within the UKB cohort was assessed using principal component analysis, with European ancestry defined by 4-means clustering on the first two genetic principal components (Warren et al., 2017). A two-stage imputation was performed using the Haplotype Reference Consortium (HRC) and UK10K reference panels (Bycroft et al., 2018; McCarthy et al., 2016; Walter et al., 2015). Poor imputed variants were excluded (Bycroft et al., 2018) (INFO ≤ 0.4).

**GSRD and HSR cohorts**: The processing and quality control pipelines applied to **replication cohorts** were implemented with similar criteria.

In GSRD, genotyping was performed using the Illumina Infinium PsychArray 24 BeadChip (Illumina, Inc., San Diego). Pre-imputation quality control was performed as previously described (PMID 30468137), i.e., for variants: missing rate ≥5% or monomorphic; and for participants: genotyping rate <97%, sex discrepancies, abnormal heterozygosity, identity by descent (IBD) >0.1875, population outliers according to Eigensoft analysis of linkage-disequilibrium-pruned genetic data. Genotypes were imputed using the Haplotype Reference Consortium (HRC) r1.1 2016 data as reference panel and Minimac3. Post-imputation quality control excluded variants with poor imputation quality (R 2<0.30) or minor allele frequency (MAF) <0.01.

In HSR, blood samples were genotyped using the Infinium PsychArray 24 BeadChip (Illumina, Inc., San Diego). QC was conducted with PLINK1.9 (Purcell et al., 2007). Individuals with discrepant genotyped sex information, genotype rate < 95%, or outlying autosomal heterozygosity (Fhet > ±0.2) were removed. Markers with minor allele frequency (MAF) < 1%, call rate < 95%, or deviant from Hardy-Weinberg equilibrium at p<10−6 were excluded. Relatedness of participants was checked by excluding individuals with a degree of recent shared ancestry (i.e., identity by descendent, IBD) > 0.1875, which is the threshold corresponding to the halfway between third- and second-degree relatives (Anderson et al., 2011). Finally, European population ancestry of the sample was confirmed with a principal component analysis (PCA), removing individuals whose genotype distribution was deviant from more than 5 standard deviations from the mean of the first two components. After QC, the Michigan Imputation Server (https://imputationserver.sph.umich.edu/index.html) was used for genotype imputation and PRS calculation. The post-QC dataset was imputed using the Haplotype Reference Consortium panel (i.e., HRC r1.1 2016) as reference panel, adopting Minimac4 and setting the *rsq* filter to 0.003.

**Supplementary Figures**

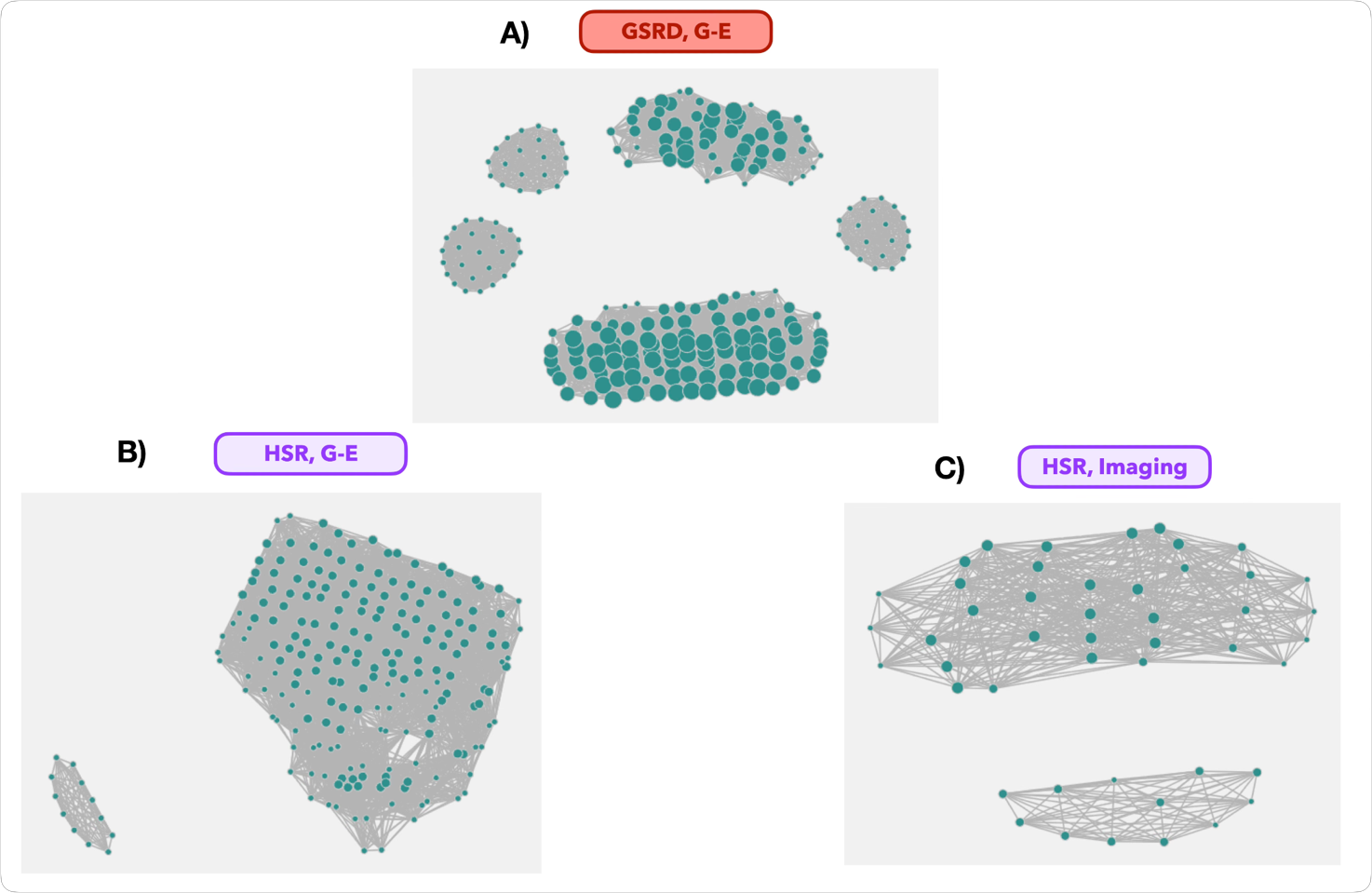

**Figure S1. TDA graph structures for UKB-derived best predictors showing significant cross-cohort replication in GSRD and HSR sets**.TDA graph structures for UKB-derived best predictors that successfully differentiated outcome subgroups in independent replication samples of GSRD and HSR: **(A)** TDA graph extracted on G-E feature set for GSRD, **(B)** TDA graph extracted on G-E feature set for HSR, **(C)** TDA graph extracted on Imaging feature set for HSR.

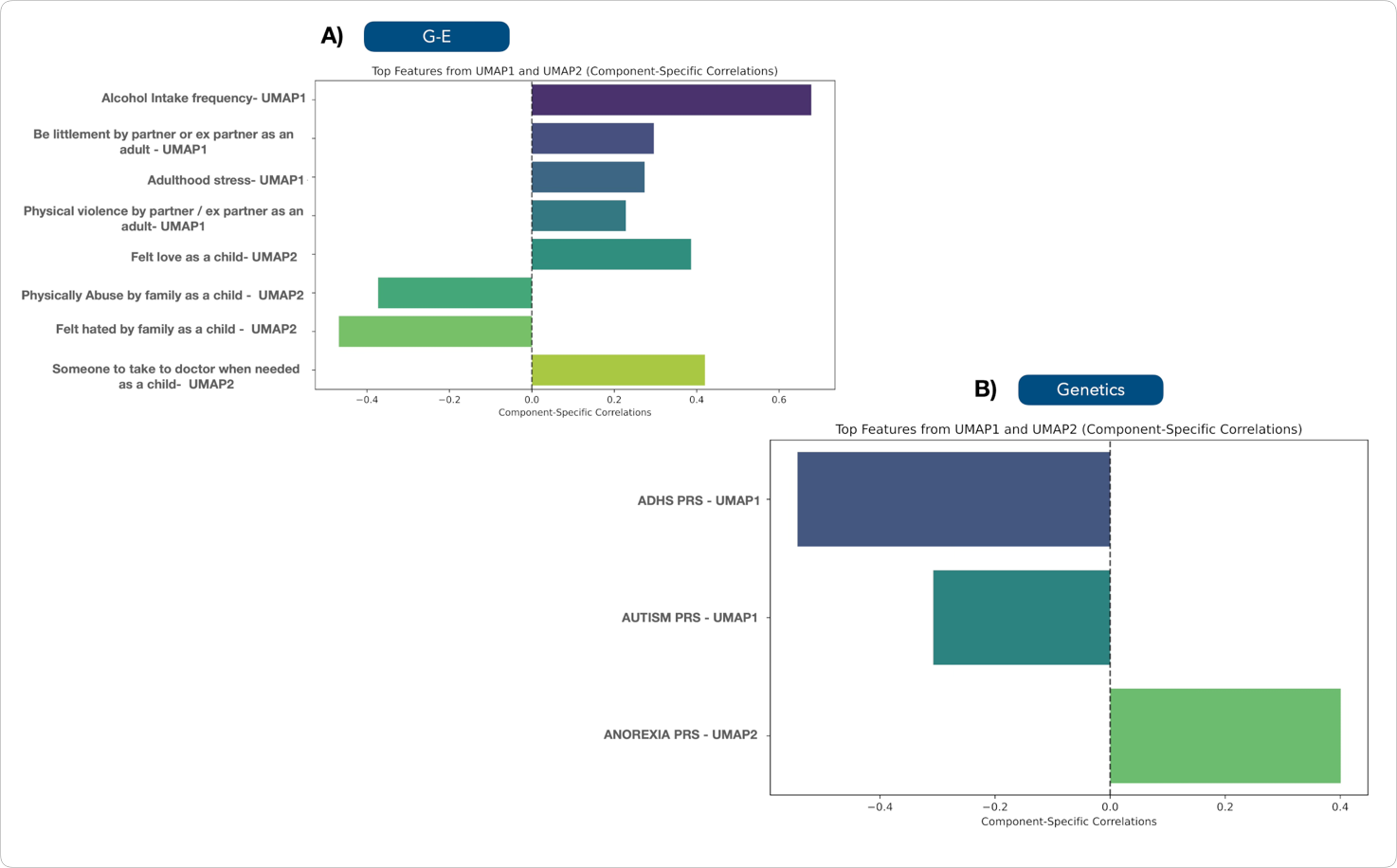

**Figure S2. UMAP feature embedding correlations for UKB best predictors of G-E and Genetics feature sets. Feature-wise correlation strengths with UMAP embedding components for G-E and Genetics best predictor.** Correlation values with UMAP1 and UMAP2 components are shown for the features of the best predictor feature sets surviving multiple comparison correction and selected for subsequent community-based validation in the two-stage feature ranking pipeline.

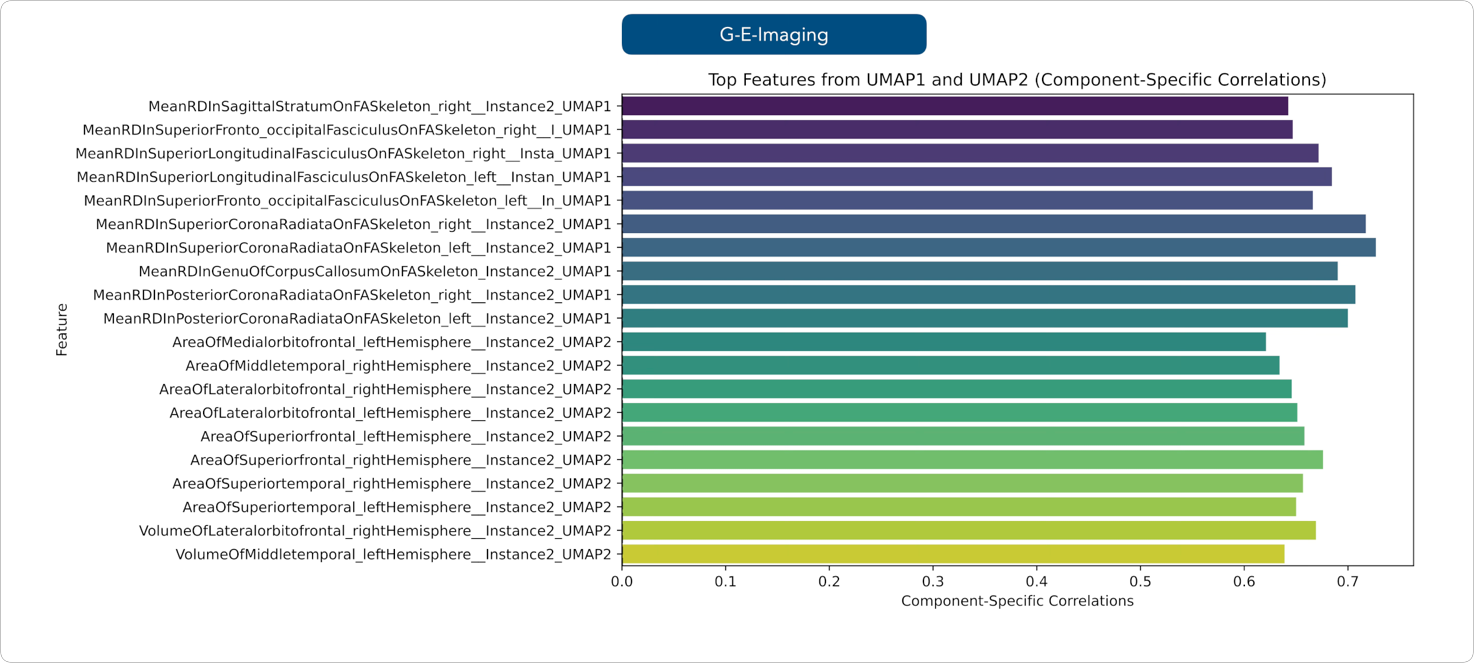
 **Figure S3**. **UMAP feature embedding correlations for UKB best predictors of G-E-Imaging feature set. Feature-wise correlation strengths with UMAP embedding components for G-E-Imaging best predictor.** Correlation values with UMAP1 and UMAP2 components are shown for the features of the best predictor feature sets surviving multiple comparison correction and selected for subsequent community-based validation in the two-stage feature ranking pipeline.

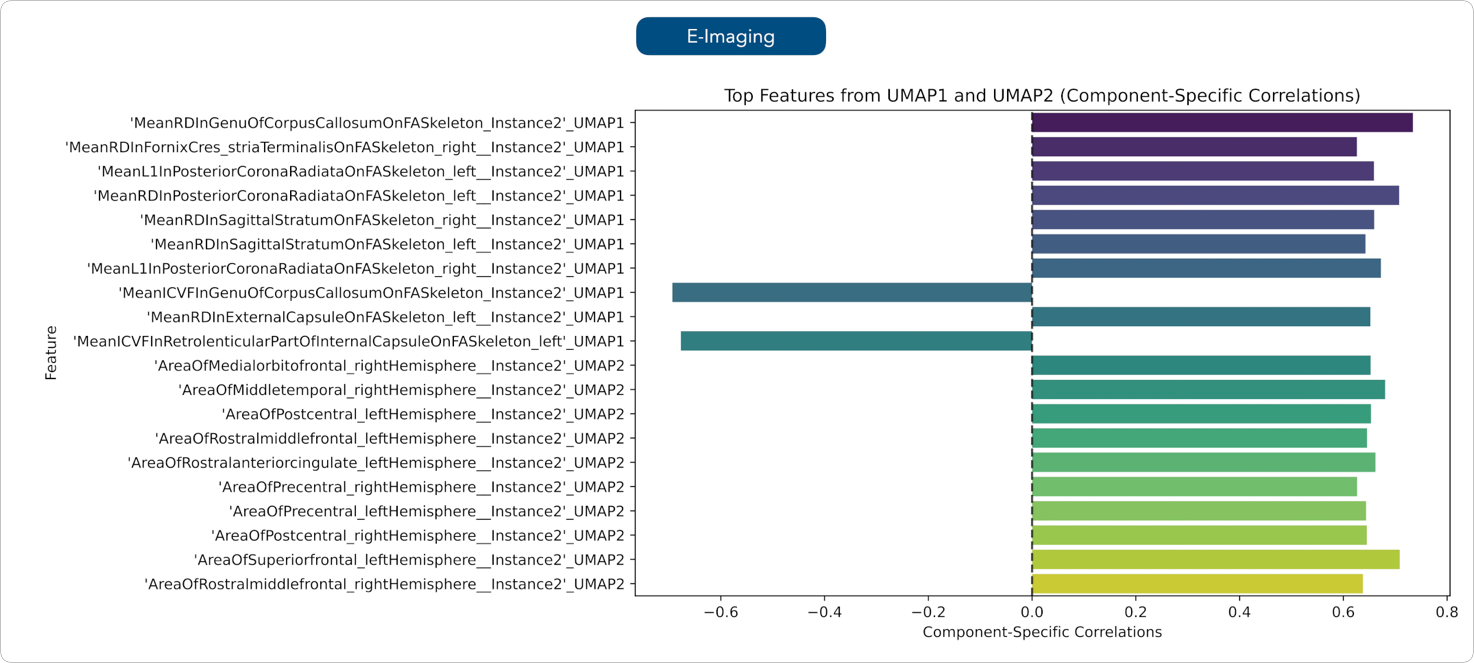

**Figure S4**. **UMAP feature embedding correlations for UKB best predictors of E-Imaging feature set**. **Feature-wise correlation strengths with UMAP embedding components for E-Imaging best predictor.** Correlation values with UMAP1 and UMAP2 components are shown for the features of the best predictor feature sets surviving multiple comparison correction and selected for subsequent community-based validation in the two-stage feature ranking pipeline.

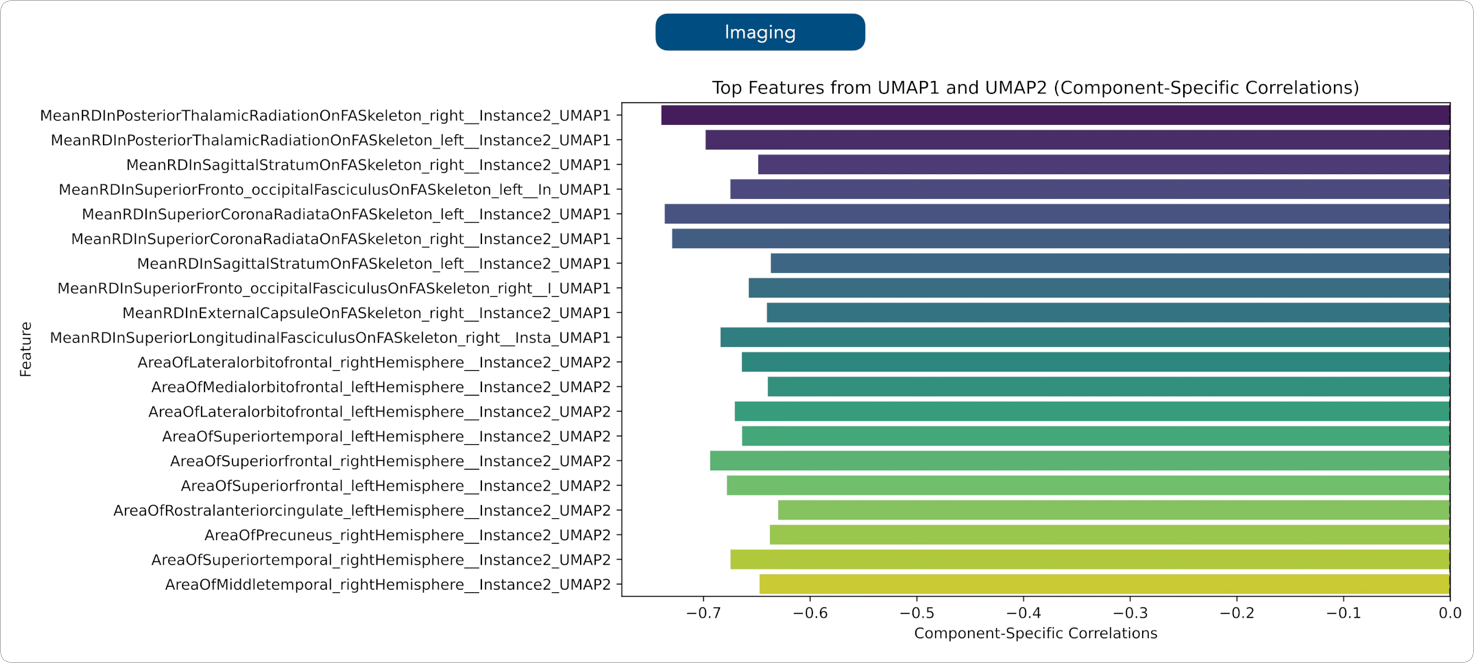

**Figure S5 UMAP feature embedding correlations for UKB best predictors of Imaging feature set. Feature-wise correlation strengths with UMAP embedding components for Imaging best predictor.** Correlation values with UMAP1 and UMAP2 components are shown for the features of the best predictor feature sets surviving multiple comparison correction and selected for subsequent community-based validation in the two-stage feature ranking pipeline.

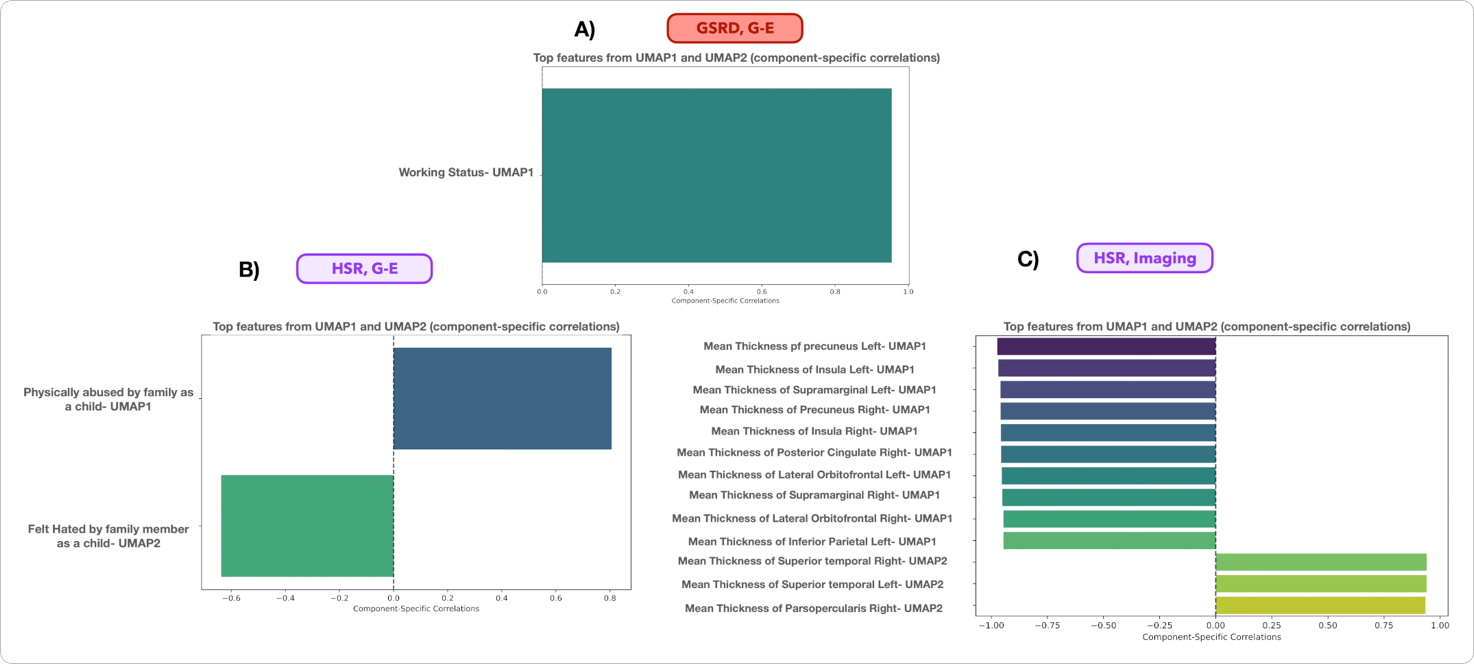

**Figure S6**. **UMAP feature embedding correlations for replication sets obtained on UKB-derived best predictors that successfully differentiated outcome subgroups in GSRD and HSR replication samples**. **Feature-wise correlation strengths with UMAP embedding components for UKB-derived best predictor showing significant cross-cohort replication.** Correlation values with UMAP1 and UMAP2 components are shown for the features of the UKB-derived best predictor feature sets surviving multiple comparison correction and selected for subsequent community-based validation in the two-stage feature ranking pipeline. **(A)** **Feature-wise correlation strengths with UMAP embedding components for G-E feature set of GSRD, (B) Feature-wise correlation strengths with UMAP embedding components for G-E feature set of HSR, (C) Feature-wise correlation strengths with UMAP embedding components for Imaging feature set of HSR.**

**Supplementary Tables**

**Table S1.** Resting-state networks associated with the 55 non-artefactual ICs generated from group-ICA of UKB cohort.

| **RSNs** | Non-artefactual ICs included |
| --- | --- |
| VAN | IC-2, -15, -37, -45, -47 |
| VIS | IC-1, -3, -16, -8, -14 |
| DMNp | IC-5, -10, -11, -19, -36 |
| SMN | IC-6, -20, -22, -30, -32, -35 |
| DMNa | IC-7, -13, -21, -28, -52, -55 |
| Sub&Cereb | IC-38, -17, -23, -53, -54 |
| DAN | IC-5, -18, -27, -34, -39, -40, -41, -42 |
| TempPar | IC-9, -33, -43, -48, -50, -51 |
| FPN | IC-12, -24, -25, -26, -29, -31, -44, -46, -49 |

Abbreviations: RSNs, resting-state networks; ICs, independent components; DMNa, default mode network anterior; DMNp, default mode network posterior; SMN, somatomotor network; Sub&Cereb, subcortical and cerebellar network; TempPar, temporoparietal network; VAN, ventral attention network; DAN, dorsal attention network; VIS, visual network ; FPN, frontoparietal network.

**Table S2.** Genome-wide association studies used to estimate the effect of genetic variants associated with various traits and further for the calculation of polygenic risk scores.

| **Trait** | **Type of trait** | **N cases** | **N controls** | **Ethnicity** | **PMID** |
| --- | --- | --- | --- | --- | --- |
| MDD | Binary | 45,591 | 97,674 | European | 29700475 |
| BD | Binary | 20,352 | 31,358 | European | 31043756 |
| Schizophrenia | Binary | 40,675 | 64,643 | European | 29483656 |
| ADHD | Binary | 20,183 | 35,191 | European | 30478444 |
| Anorexia nervosa | Binary | 16,992 | 55,525 | European | 31308545 |
| Autism | Binary | 18,381 | 27,969 | European | 30804558 |
| Alcohol Dependence | Binary | 14,904 | 37,944 | Mixed | 30482948 |
| Daily alcohol consumption | Continuous | 70,460 | | European | 27911795 |
| BMI | Continuous | 322,154 | | European | 25673413 |
| Coronary artery disease | Binary | 60,801 | 123,504 | European e Asiatic | 26343387 |
| C reactive protein | Binary | 204,402 | | European | 30388399 |
| Type 2 diabetes mellitus | Binary | 26,676 | 132,532 | Mixed | 28566273 |
| Glycated hemoglobin | Continuous | 123,665 | | European | 28898252 |
| N sigarette / day | Continuous | 38,181 | | European | 20418890 |
| Triglycerides | Continuous | 1,654,960 | | Mixed | 34887591 |
| LDL cholesterol | Continuous | 1,654,960 | | Mixed | 34887591 |
| HDL cholesterol | Continuous | 1,654,960 | | Mixed | 34887591 |
| Years of education | Continuous | 766,345 | | European | 30038396 |
| Neuroticism | Continuous | 63,661 | | European | 25993607 |

Abbreviations: Bipolar disorder (BD-PRS), schizophrenia (SCZ-PRS), MDD (DEPR07-PRS), anorexia nervosa (ANOR07-PRS), autism (AUTI07-PRS), attention deficit hyperactivity disorder (ADHD05-PRS). PRS related to the following non-psychiatric traits were extracted: smoking (SMOK03-PRS), alcohol dependence (ALCD01_PRS), C reactive protein (CRP-PRS), years of education (education-PRS), glycated hemoglobin (hb1ac-PRS), triglycerides (TRGL_PRS), LDL cholesterol (LDL-PRS), coronary artery disease (CAD-PRS), alcohol consumption (ALCO01_PRS), body mass index (BMI-PRS), HDL cholesterol (HDL_PRS), type 2 diabetes mellitus (DM-PRS), neuroticism (Neuroticism-PRS).

**Table S3.** Environmental characteristics included and relative UKB code.

| **Environmental Set** | | |  |
| --- | --- | --- | --- |
| Name | UKB Data field or definition | Variable Domain original | Variable Domain modified |
| Frequency of friend / family visits | [1031](https://biobank.ndph.ox.ac.uk/showcase/field.cgi?id=1031) | 7 levels: 1 almost every day, 2 2-4 times a week, 3 about once a week, 4 about once a month, 5 once every few months, 6 never or almost never, 7 no family/friends outside household | Binary: 0 no frequent visit (including once every few months, never or almost never, no family/friends outside household), 1 frequent visit (including almost daily, 2-4 times a week, about once a week, about once a month). |
| Leisure / social activities | [6160](https://biobank.ndph.ox.ac.uk/showcase/field.cgi?id=6160) | 5 levels: 1 Sports clubs or gym, 2 pub or social club, 3 religious group, 4 adult education class, 5 other group of activity | Binary: 0 never, 1 any type of leisure/social activities (including: sports clubs or gym, pub or social club, religious group, adult education class, other group of activity) |
| Able to confide | [2110](https://biobank.ndph.ox.ac.uk/showcase/field.cgi?id=2110) | 6 levels: 5 almost every day, 4 2.4 times a week, 3 about once a week, 2 about once a month, 1 once every few months, 0 never or almost never. | Binary: 0 lack of social support (including: 2.4 times a week, about once a week, about once a month, once every few months, never or almost never), 1 regular social support ( almost every day) |
| Morning/evening person (chronotype) | [1180](https://biobank.ndph.ox.ac.uk/showcase/field.cgi?id=1180) | 4 levels: 1 definitely a “morning person”, 2 more a “morning person” then “evening person”, 3 more an “evening person” than a “morning person”, 4 definitely an “evening person”. | Binary: 0 morning person (including: more an “evening person” than a “morning person”, definitely an “evening person”), 1 evening person (including: definitely a “morning person”, more a “morning person” then “evening person”) |
| Summed MET minutes per week for all activity | [22040](https://biobank.ndph.ox.ac.uk/showcase/field.cgi?id=22040) | Continuous | / |
| Z adjusted T/S log  (i.e., Telomeres length) | [22192](https://biobank.ndph.ox.ac.uk/showcase/field.cgi?id=22192) | Continuous | / |
| Neuroticism-EPQ RS | Derived from [10.1038/mp.2016.49](https://doi.org/10.1038%2Fmp.2016.49) | / | 12 levels extracted from 12 items of neuroticism scale from the Eysenck Personality Questionnaire-Revised Short Form. |
| Felt love as a child | [20489](https://biobank.ndph.ox.ac.uk/showcase/field.cgi?id=20489) | 5 levels: 0 never, 1 rarely true, 2 sometimes true, 3 often, 4 very often | / |
| Physically abused by family as a child | [20488](https://biobank.ndph.ox.ac.uk/showcase/field.cgi?id=20488) | 5 levels: 0 never, 1 rarely true, 2 sometimes true, 3 often, 4 very often | / |
| Felt hated by family member as a child | [20487](https://biobank.ndph.ox.ac.uk/showcase/field.cgi?id=20487) | 5 levels: 0 never, 1 rarely true, 2 sometimes true, 3 often, 4 very often | / |
| Sexually molested as a child | [20490](https://biobank.ndph.ox.ac.uk/showcase/field.cgi?id=20490) | 5 levels: 0 never, 1 rarely true, 2 sometimes true, 3 often, 4 very often | / |
| Someone to take to doctor when needed as a child | [20491](https://biobank.ndph.ox.ac.uk/showcase/field.cgi?id=20491) | 5 levels: 0 never, 1 rarely true, 2 sometimes true, 3 often, 4 very often | / |
| Been in a confiding relationship as an adult | [20522](https://biobank.ndph.ox.ac.uk/showcase/field.cgi?id=20522) | 5 levels: 0 never, 1 rarely true, 2 sometimes true, 3 often, 4 very often | / |
| Physical violence by partner or ex-partner as an adult | [20523](https://biobank.ndph.ox.ac.uk/showcase/field.cgi?id=20523) | 5 levels: 0 never, 1 rarely true, 2 sometimes true, 3 often, 4 very often | / |
| Belittlement by partner or ex-partner as an adult | [20521](https://biobank.ndph.ox.ac.uk/showcase/field.cgi?id=20521) | 5 levels: 0 never, 1 rarely true, 2 sometimes true, 3 often, 4 very often | / |
| Sexual interference by partner or ex-partner without consent as an adult | [20524](https://biobank.ndph.ox.ac.uk/showcase/field.cgi?id=20524) | 5 levels: 0 never, 1 rarely true, 2 sometimes true, 3 often, 4 very often | / |
| Able to pay rent/mortgage as an adult | [20525](https://biobank.ndph.ox.ac.uk/showcase/field.cgi?id=20525) | 5 levels: 0 never, 1 rarely true, 2 sometimes true, 3 often, 4 very often | / |
| Victim of sexual assault | [20531](https://biobank.ndph.ox.ac.uk/showcase/field.cgi?id=20531) | 3 levels: 0 never, 1 yes but not in the last 12 months, 2 yes within the last 12 months. | Binary: 0 never, 1 it happened (including: yes but not in the last 12 months, yes within the last 12 months) |
| Victim of physically violent crime | [20529](https://biobank.ndph.ox.ac.uk/showcase/field.cgi?id=20529) | 3 levels: 0 never, 1 yes but not in the last 12 months, 2 yes within the last 12 months. | Binary: 0 never, 1 it happened (including: yes but not in the last 12 months, yes within the last 12 months) |
| Been in serious accident believed to be life-threatening | [20526](https://biobank.ndph.ox.ac.uk/showcase/field.cgi?id=20526) | 3 levels: 0 never, 1 yes but not in the last 12 months, 2 yes within the last 12 months. | Binary: 0 never, 1 it happened (including: yes but not in the last 12 months, yes within the last 12 months) |
| Witnessed sudden violent death | [20530](https://biobank.ndph.ox.ac.uk/showcase/field.cgi?id=20530) | 3 levels: 0 never, 1 yes but not in the last 12 months, 2 yes within the last 12 months. | Binary: 0 never, 1 it happened (including: yes but not in the last 12 months, yes within the last 12 months) |
| Diagnosed with life-threatening illness | [20528](https://biobank.ndph.ox.ac.uk/showcase/field.cgi?id=20528) | 3 levels: 0 never, 1 yes but not in the last 12 months, 2 yes within the last 12 months. | Binary: 0 never, 1 it happened (including: yes but not in the last 12 months, yes within the last 12 months) |
| Been involved in combat or exposed to war-zone | [20527](https://biobank.ndph.ox.ac.uk/showcase/field.cgi?id=20527) | 3 levels: 0 never, 1 yes but not in the last 12 months, 2 yes within the last 12 months. | Binary: 0 never, 1 it happened (including: yes but not in the last 12 months, yes within the last 12 months) |
| Alcohol intake frequency | [1558](https://biobank.ndph.ox.ac.uk/showcase/field.cgi?id=1558) | 6 levels: 1 daily or almost daily, 2 three or four times a week, 3 once or twice a week, 4 once to three times a month, 5 special occasions only, 6 never. | 4 levels: 0 never, 1 special occasion only or sometimes a month, 2 sometimes a week, 3 almost daily |
| Childhood adverse events | Derived from doi: 10.1016/j.ynstr.2022.100447 based on: [20489](https://biobank.ndph.ox.ac.uk/showcase/field.cgi?id=20489),  [20488](https://biobank.ndph.ox.ac.uk/showcase/field.cgi?id=20488), [20487](https://biobank.ndph.ox.ac.uk/showcase/field.cgi?id=20487), [20490](https://biobank.ndph.ox.ac.uk/showcase/field.cgi?id=20490), [20491](https://biobank.ndph.ox.ac.uk/showcase/field.cgi?id=20491) | / | 5 levels. |
| Adulthood stress | Derived from 10.1016/j.ynstr.2022.100447 based on:  [20487](https://biobank.ndph.ox.ac.uk/showcase/field.cgi?id=20487), [20488](https://biobank.ndph.ox.ac.uk/showcase/field.cgi?id=20488), [20489](https://biobank.ndph.ox.ac.uk/showcase/field.cgi?id=20489), [20490](https://biobank.ndph.ox.ac.uk/showcase/field.cgi?id=20490), [20491](https://biobank.ndph.ox.ac.uk/showcase/field.cgi?id=20491) | / | Continous variable |
| Serious illness, injury, assault to yourself or assault of a close relative in the last 2 years | [6145](https://biobank.ndph.ox.ac.uk/showcase/field.cgi?id=6145) | 6 levels: 1 serious illness, injury or assault to yourself, 2 serious illness, injury or assault of a close relative, 3 death of a close relative, 4 death of a spouse or partner, 5 marital separation/divorce, 6 financial difficulties | Binary: 1 if it happened (including: serious illness, injury or assault to yourself, serious illness, injury or assault of a close relative ), 0 None (including all the other categories). |
| Death of a close relative or death of a spouse or partner in the last 2 years | [6145](https://biobank.ndph.ox.ac.uk/showcase/field.cgi?id=6145) | 6 levels: 1 serious illness, injury or assault to yourself, 2 serious illness, injury or assault of a close relative, 3 death of a close relative, 4 death of a spouse or partner, 5 marital separation/divorce, 6 financial difficulties | Binary: 1 if it happened (including: death of a close relative, death of a spouse or partner), 0 None ( including all the other categories), |
| Stress related to marital separation/divorce or to financial difficulties in the last 2 years | [6145](https://biobank.ndph.ox.ac.uk/showcase/field.cgi?id=6145) | 6 levels: 1 serious illness, injury or assault to yourself, 2 serious illness, injury or assault of a close relative, 3 death of a close relative, 4 death of a spouse or partner, 5 marital separation/divorce, 6 financial difficulties | Binary: 1 if it happened (including: marital separation/divorce, financial difficulties), 0 None ( including all the other categories). |
| Ever taken cannabis | [20453](https://biobank.ndph.ox.ac.uk/showcase/field.cgi?id=20453) | 5 levels: 0 no, 1 yes 1-2 times, 2 yes 3-10 times, 3 yes 11-100 times, 4 yes more than 100 times. | 3 levels: 0 never, 1 if at maximum 10 times, 2 if more than 10 times |
| Smoking status | [20116](https://biobank.ndph.ox.ac.uk/showcase/field.cgi?id=20116) | 3 levels: 0 if never, 1 if previous, 2 if current | / |
| Major dietary changes because of illness in the last 5 years | [1538](https://biobank.ndph.ox.ac.uk/showcase/field.cgi?id=1538) | 3 levels: 0 no, 1 yes because of illness, 2 yes because of other reasons. | Binary: 0 never, 1 if it happened. |
| Major dietary changes because other reason in the last 5 years | [1538](https://biobank.ndph.ox.ac.uk/showcase/field.cgi?id=1538) | 3 levels: 0 no, 1 yes because of illness, 2 yes because of other reasons. | Binary: 0 never, 1 if it happened. |

Abbreviations**:** MET, metabolic equivalent task.

**Table S4**. List of socio-demographic characteristics of the sample included with the relative UKB code.

| UKB Data field or definition | Name |
| --- | --- |
| [21003](https://biobank.ndph.ox.ac.uk/showcase/field.cgi?tk=yZjRoJFA3dq4WUv7alKluuMBHS4bpDM7189922&id=21003)  [31](https://biobank.ndph.ox.ac.uk/showcase/field.cgi?tk=yZjRoJFA3dq4WUv7alKluuMBHS4bpDM7189922&id=31)  [21000](https://biobank.ndph.ox.ac.uk/showcase/field.cgi?tk=yZjRoJFA3dq4WUv7alKluuMBHS4bpDM7189922&id=21000)  [21001](https://biobank.ndph.ox.ac.uk/showcase/field.cgi?tk=yZjRoJFA3dq4WUv7alKluuMBHS4bpDM7189922&id=21001)  [48](https://biobank.ndph.ox.ac.uk/showcase/field.cgi?tk=yZjRoJFA3dq4WUv7alKluuMBHS4bpDM7189922&id=48) | Age at baseline  Sex  Ethnicity  BMI  WC |

Abbreviations: BMI, Body mass index; WC, waist circumference.

**Table S5**. List of drugs included in antidepressant medications with the relative UKB code.

| UKB Data field or definition | Name |
| --- | --- |
| 1140879616  1140921600  1140879540  1140867878  1140916282  1140909806  1140867888  1141152732  1141180212  1140879634  1140867876  1140882236  1141190158  1141200564  1140867726  1140879620  1140867818  1140879630  1140879628  1141151946  1140867624  1140867756  1140867884  1141151978  1141152736  1141201834  1140867690  1140867640  1140867920  1140867850  1140879544  1141200570  1140867934  1140867758  1140867914  1140867820  1141151982  1140882244  1140879556  1140867852  1140867860  1140917460  1140867938  1140867856  1140867922  1140910820  1140882312  1140867944  1140867784  1140867812  1140867668  1140867940  1140867948  1140867774  1141174756  1140916288  1140867628  1140882310  1140867784  1140867930  1140867712  1140867916 | amitriptyline  citalopram  fluoxetine  sertraline  venlafaxine  dosulepin  paroxetine  mirtazapine  escitalopram  trazodone  prozac  seroxat  cipralex  duloxetine  lofepramine  clomipramine  nortriptyline  imipramine  dothiepin  cipramil  prothiaden  trimipramine  lustral  reboxetine  zispin  cymbalta  anafranil  doxepin  moclobemide  phenelzine  fluvoxamine  yentreve  triptafen  surmontil  tranylcypromine  allegron  edronax  molipaxin  mianserin  nardil  faverin  nefazodone  amitriptyline+chlordiazepoxide  isocarboxazid  manerix  maoi  sinequan  tranylcypromine+trifluoperazine  ludiomil  norval  tryptizol  fluphenazine hydrochloride+nortriptyline  amitriptyline hydrochloride+perphenazine 10mg/2mg tablet  amoxapine  oxactin 20mg capsule  efexor 37.5mg tablet  prepadine 25mg capsule  gamanil 70mg tablet  ludiomil  motival  tofranil  parnate |

**Table S6**. List of antidepressant mnedications in HSR cohort

List of antidepressant medications

Venlafaxine

Escitalopram

Citalopram

Sertraline

Mirtazapine

Trazodone

Fluvoxamine

Fluoxetine

Duloxetine

Paroxetine

Clomipramine

Imipramine

Nortriptyline

Trimipramine

Vortioxetine

Bupropion

Amitriptyline

**Table S7**. Outcomes included in each group with the relative UKB code.

| **Group 1 (G1)** | | | | |
| --- | --- | --- | --- | --- |
| Name | UKB Data field or definition | Variable Domain original | Variable Domain modified | |
| O1: Duration of worst depression | [20438](https://biobank.ndph.ox.ac.uk/showcase/field.cgi?id=20438) | 3 levels: 1=1-2 Below 3 months, 2=3-4 below 1 year=2, 3=5,6 up to one year |  | |
| O2: Frequency of depressed days during worst episode of depression | [20439](https://biobank.ndph.ox.ac.uk/showcase/field.cgi?id=20439) | 3 levels: 1= less often, 2 = almost every day, 3= every day |  | |
| O3: Impact on normal roles during worst period of depression | [20440](https://biobank.ndph.ox.ac.uk/showcase/field.cgi?id=20440) | 3 levels: 0 not at all, 1 a little, 2 somewhat, 3 a lot |  | |
| O4: Thoughts of death during worst depression | [20437](https://biobank.ndph.ox.ac.uk/showcase/field.cgi?id=20437) | Binary: 0 none, 1 if it happened |  | |
| O5: Ever self harmed | [20480](https://biobank.ndph.ox.ac.uk/showcase/field.cgi?id=20480) | Binary: 0 none, 1 if it happened |  | |
| O6: Belief that owns life is meaningful | [20460](https://biobank.ndph.ox.ac.uk/showcase/field.cgi?id=20460) | 5 levels: 1 Not at all, 2 A little, 3 A moderate amount, 4 Very much, 5 An extreme amount |  | |
| O7: Depression possibly related to stressful or traumatic event | [20447](https://www.google.com/url?q=https://biobank.ndph.ox.ac.uk/showcase/field.cgi?tk%3DyZjRoJFA3dq4WUv7alKluuMBHS4bpDM7189922%26id%3D20447&sa=D&source=editors&ust=1643042170960687&usg=AOvVaw3_ixJzwm70S9U7QYtNZuDB) | Binary: 0 condition not present, 1 condition present |  | |
| O8: Depression with atypical neurovegetative symptoms | Derived from <https://doi.org/10.1038%2Fs41386-021-01059-6>. | Binary: 0 condition not present, 1 condition present |  | |
| O9: Depression with anxious features | Derived from <https://doi.org/10.1038%2Fs41386-021-01059-6>. | Binary: 0 condition not present, 1 condition present |  | |
| **Group 2 (G2)** | | | | |
| Name | UKB Data field or definition | Variable Domain original | | Variable Domain modified |
| O1: Cancer diagnosed by doctor | [2453](https://biobank.ndph.ox.ac.uk/showcase/field.cgi?id=2453) | Binary: 0 not diagnosed, 1 diagnosed | | / |
| O2: Vascular heart problems | [6150](https://biobank.ndph.ox.ac.uk/showcase/field.cgi?id=6150) | 3 levels: 2 if HBP, heart attack, stroke or angina, 1 if HBP, 0 if none | | 3 levels: 0=None, 1=HBP, 2=HBP with Stroke, or Angina or Heart Attack |
| O3: Diabetes diagnosed by doctor | [2443](https://biobank.ndph.ox.ac.uk/showcase/field.cgi?id=2443) | Binary: 0 not diagnosed, 1 diagnosed | | / |
| **Group 3 (G3)** | | | | |
| O1: Treatment resistant depression | Derived from [10.1038/s41380-021-01062-9](https://doi.org/10.1038/s41380-021-01062-9). | / | | Binary: 0 condition not present, 1 condition present |

Abbreviations: G, group of clinical outcomes; O, clinical outcome.

**Table S8.** List of genetics, environmental and socio-demographic characteristics included for the GSRD and HSR independent sets, with the corresponding UKB code and domain definition.

| **Genetic, Environmental and Socio-demographic variables***  ********In case of similar but not perfect matching with UKB, variables were derived from anamnesis or clinical questionnaires, such as Childhood Trauma Questionnaire (CTQ), Schedule of Recent Experience (SRE), 13-items Beck Depression Inventory (BDI), or the World Health Organization Quality of Life.* | | | | | | | | |
| --- | --- | --- | --- | --- | --- | --- | --- | --- |
| **Genetic variables (PRS)** | | | | | | | | |
| **GSRD (P=18)** | | | | | | **HSR (P=19)** | | |
| BD  Schizophrenia  MDD  Anorexia Nervosa  Autism  ADHD  Alcohol dependece  Smoking (N sigarette / day)  Alcohol consumption (daily)  Years of education  C-reactive protein  Glycated hemoglobin  Triglycerides  LDL cholesterol  HDL cholesterol  Coronary artery disease  BMI  Type 2 diabetes mellitus | | | | | | BD  Schizophrenia  MDD  Anorexia Nervosa  Autism  ADHD  Alcohol dependece  Neuroticism  Smoking (N sigarette / day)  Alcohol consumption (daily)  Years of education  C-reactive protein  Glycated hemoglobin  Triglycerides  LDL cholesterol  HDL cholesterol  Coronary artery disease  BMI  Type 2 diabetes mellitus | | |
| **Environment variables** | | | | | | | | |
| **UKB**  **Code \| Variable domain definition** | | | **GSRD (P=5)** | | | **HSR (P=12)** | | |
|  |  |  | **Name** | | **Variable domain definition** | **Name** | | **Variable domain definition** |
| Leisure / social activities, 6160 | Definition: Binary: 0 never, 1 any type of leisure/social activities (including: sports clubs or gym, pub or social club, religious group, adult education class, other group of activity) | | Leisure / social activities | Definition: Sheenan Disability Scale, item social activities, scores 1-6 vs 7-10 item | | / | / | |
| Been in a confiding relationship as an adult, 20522 | Definition: 5 levels: 0 never, 1 rarely true, 2 sometimes true, 3 often, 4 very often | | Marital Status | Definition: Marital Status, Binary: 0: Single, 1: with partner | | / | / | |
| Able to pay rent/mortgage as an adult, 20525 | Definition: 5 levels: 0 never, 1 rarely true, 2 sometimes true, 3 often, 4 very often | | Working Status | Definition: Working Status, Binary: 0: Unemployed, 1: Working | | / | / | |
| Alcohol intake frequency, 1558 | Definition: 4 levels: 0 never, 1 special occasion only or sometimes a month, 2 sometimes a week, 3 almost daily | | Alcohol intake frequency | Definition**:** Any Abuse anamnesis, Binary: 0: None, 1: if it happened | | Alcohol intake frequency | Domain definition: Anamnesis, Binary: 0: None, 1: special occasions only, 2: sometimes/week, 3: almost every day | |
| Smoking status, 20116 | Definition: 3 levels: 0 if never, 1 if previous, 2 if current | | Smoking status | Definition: Anamnesis, Binary: 0: No, 1: Yes | | Smoking status | Domain definition: 3 levels: 0 if never, 1 if previous or current | |
| Felt love as a child, [20489](https://biobank.ndph.ox.ac.uk/showcase/field.cgi?id=20489) | Definition: 5 levels: 0 never, 1 rarely true, 2 sometimes true, 3 often, 4 very often | | / | / | | Felt love as a child | Definition: CTQ item 7 | |
| Physically abused by family as a child, [20488](https://biobank.ndph.ox.ac.uk/showcase/field.cgi?id=20488) | Definition: 5 levels: 0 never, 1 rarely true, 2 sometimes true, 3 often, 4 very often | | / | / | | Physically abused by family as a child | Definition: CTQ item 11 | |
| Felt hated by family member as a child, [20487](https://biobank.ndph.ox.ac.uk/showcase/field.cgi?id=20487) | Definition: 5 levels: 0 never, 1 rarely true, 2 sometimes true, 3 often, 4 very often | | / | / | | Felt hated by family member as a child | Definition: CTQ item 18 | |
| Sexually molested as a child, [20490](https://biobank.ndph.ox.ac.uk/showcase/field.cgi?id=20490) | Definition: 5 levels: 0 never, 1 rarely true, 2 sometimes true, 3 often, 4 very often | | / | / | | Sexually molested as a child | Definition: CTQ item 24 | |
| Someone to take to doctor when needed as a child, [20491](https://biobank.ndph.ox.ac.uk/showcase/field.cgi?id=20491) | Definition: 5 levels: 0 never, 1 rarely true, 2 sometimes true, 3 often, 4 very often | | / | / | | Someone to take to doctor when needed as a child | Definition: CTQ item 26 | |
| Childhood adverse events, derived from doi: 10.1016/j.ynstr.2022.100447 | Definition: 5 levels. | | / | / | | Childhood adverse events | Definition: Sum of CTQ items7 (reverse), CTQ item 11, CTQ item 18, CTQ item 24, CTQ item 26 (reverse) | |
| Serious illness, injury, assault to yourself or assault of a close relative in the last 2 years, [6145](https://biobank.ndph.ox.ac.uk/showcase/field.cgi?id=6145) | Definition: Binary: 1 if it happened (including: serious illness, injury or assault to yourself, serious illness, injury or assault of a close relative ), 0 None (including all the other categories). | | / | / | | Serious illness, injury, assault to yourself or assault of a close relative in the last 3 years | Definition: SRE 3 year, Binary: 0 if items 13 (accident) and 18 (illness/injury) over 3-year period are both 0, otherwise 1. | |
| Death of a close relative or death of a spouse or partner in the last 2 years, [6145](https://biobank.ndph.ox.ac.uk/showcase/field.cgi?id=6145) | Definition: Binary: 1 if it happened (including: death of a close relative, death of a spouse or partner), 0 None ( including all the other categories) | | / | / | | Death of a close relative or death of a spouse or partner in the last 3 years | Definition: SRE 3 year, Binary: 0 if items 14 (death of family member) and 15 (death of spouse) over 3-year period are both 0, otherwise 1. | |
| Stress related to marital separation/divorce or to financial difficulties in the last 2 years, [6145](https://biobank.ndph.ox.ac.uk/showcase/field.cgi?id=6145) | Definition: Binary: 1 if it happened (including: marital separation/divorce, financial difficulties), 0 None ( including all the other categories). | | / | / | | Stress related to marital separation/divorce or to financial difficulties in the last 3 years | Definition: SRE 3 year, Binary: 0 if items 24 (divorce) and 25 (separation) over 3-year period are both 0, otherwise 1. | |
| Major dietary changes because other reason in the last 5 years, [1538](https://biobank.ndph.ox.ac.uk/showcase/field.cgi?id=1538) | Definition: Binary: 0 never, 1 if it happened. | | / | / | | Major dietary changes because other reason in the last 5 years | Definition: SRE 3years item 3, Binary: 0 None, 1 If it happened | |
| **Confounding variables** | | | | | | | | |
| **UKB**  **Code \| Variable domain definition** | | | **GSRD (P=2)**  **Variable domain definition** | | | **HSR (P=4)**  **Variable domain definition** | | |
| **Age,** 21003 | |  | **Age** | | | **Age** | | |
| **Sex,** 31 | |  | **Sex** | | | **Sex** | | |
| **Ethnicity,** [21000](https://biobank.ndph.ox.ac.uk/showcase/field.cgi?tk=yZjRoJFA3dq4WUv7alKluuMBHS4bpDM7189922&id=21000) | | Definition: dichotomics, 0-1 (1, Caucasian, 0 others) |  | | | **Ethnicity**  Definition: Dichotomics, 0-1 (1, Caucasian, 0 others) | | |
| **Antidepressant Medication** | | Definition**:** Dichotomics, 0-1 (0=no, 1= antidepressants, from UKB codes for the list of drugs included S5) |  |  |  | **Antidepressant Medication**  Definition: Dichotomics, 0-1 (0=no, 1= antidepressants, from HSR codes for the list of drugs included S6 | | |

P, number of features; Childhood Trauma Questionnaire (CTQ) (Bernstein et al., 2003), Schedule of Recent Experience (SRE) (Holmes et al., 1967), 13-items Beck Depression Inventory (BDI) (Reynolds et al., 1981), or the World Health Organization Quality of Life (WHOQOL; PRS, Polygenetic risk scores; BD, Bipolar disorder; MDD, Major depressive disorder; ADHD, attention deficit hyperactivity disorder; LDL, Low-Density Lipoprotein cholesterol; BMI, body mass index; HDL, High-Density Lipoprotein cholesterol.

**Table S9.** List of health-related outcomes included within the GSRD and HSR independent sets with the relative UKB code if equal, or the domain definition is similar.

| **Heath-related outcomes**  ********In case of similar but not perfect matching with UKB, variables were derived from anamnesis or clinical questionnaires, such as* ***Montgomery-Asberg Depression Rating Scale (MADRS)****, Diagnostic and Statistical Manual of Mental Disorders (DSM); Mini International Neuropsychiatric Interview (MINI), 13-items Beck Depression Inventory (BDI), or the World Health Organization Quality of Life.* | | | | | |
| --- | --- | --- | --- | --- | --- |
| **UKB**  **Code \| Variable domain definition** | | **GSRD (O=5)** | | **HSR (O=10)** | |
|  |  | **Name** | **Variable domain definition** | **Name** | **Variable domain definition** |
| TRD | Definition: derived from [10.1038/s41380-021-01062-9](https://doi.org/10.1038/s41380-021-01062-9), Binary: 0 condition not present, 1 condition present | TRD | Definition: MADRS decrease <50% and >20 after two adequate treatments = TRD | TRD | Definition: Derived from Thase et al., 1997 |
| Duration of worst depression, [20438](https://biobank.ndph.ox.ac.uk/showcase/field.cgi?id=20438) | Definition: 3 levels: 1=1-2 Below 3 months, 2=3-4 below 1 year=2, 3=5,6 up to one year | Duration of current depressive episode | Definition: MINI > 365 days | Duration of current depressive episode | Definition: duration of the current depressive episode, 3 levels: 1= less than 3 months, 2=less than1 year, 3= more than one year |
| Thoughts of death during worst depression, [20437](https://biobank.ndph.ox.ac.uk/showcase/field.cgi?id=20437) | Definition: Binary: 0 none, 1 if it happened | Thoughts of death during current depressive episodes | Definition: Suicidal thoughts according to MADRS item 10 at episode onset, scores ≥4 | Thoughts of death during current depressive episodes | Definition: BDI item 8 (level 2), Binary: 0 none, 1 if it happened |
| Ever self harmed, 20480 | Definition: Binary: 0 none, 1 if it happened | Ever self harmed | Definition: Suicidal risk=High | Ever self harmed | Definition: Anamnesis, Binary: 0 none, 1 if it happened |
| Depression with anxious features, [20447](https://www.google.com/url?q=https://biobank.ndph.ox.ac.uk/showcase/field.cgi?tk%3DyZjRoJFA3dq4WUv7alKluuMBHS4bpDM7189922%26id%3D20447&sa=D&source=editors&ust=1643042170960687&usg=AOvVaw3_ixJzwm70S9U7QYtNZuDB) | Definition: derived from <https://doi.org/10.1038%2Fs41386-021-01059-6> | Depression with anxious features | Definition: Inner tension according to MADRS item 3 at episode onset, scores ≥4 | Depression with anxious features | Definition: Anamnesis DSM, Binary: 0 condition not present, 1 condition present |
| Depression with atypical neurovegetative symptoms | Definition: derived from <https://doi.org/10.1038%2Fs41386-021-01059-6>, Binary: 0 condition not present, 1 condition present | / | / | Depression with atypical neurovegetative symptoms | Definition: Anamnesis DSM-5 specifier, Binary: 0 condition not present, 1 condition present |
| Cancer diagnosed, [2453](https://biobank.ndph.ox.ac.uk/showcase/field.cgi?id=2453) | Definition: Organic comorbidities, Binary: 0 not diagnosed, 1 diagnosed | Somatic disorders (i.e., cancer, diabetes, cardiovascular disease) | **Definition:** From anamnestic clinical interview | Cancer diagnosed | Definition: Organic comorbidities, Binary: 0 not diagnosed, 1 diagnosed |
| Vascular problem diagnosed, [6150](https://biobank.ndph.ox.ac.uk/showcase/field.cgi?id=6150) | Definition: 3 levels: 0=None, 1=HBP, 2=HBP with Stroke, or Angina or Heart Attack |  |  | Vascular problems diagnosed | Definition: Organic comorbidities, Binary: 0 not diagnosed, 1 diagnosed |
| Diabetes diagnosed, [2443](https://biobank.ndph.ox.ac.uk/showcase/field.cgi?id=2443), | Definition: Binary: 0 not diagnosed, 1 diagnosed |  |  | Diabetes diagnosed | Definition: Organic comorbidities, Binary: 0 not diagnosed, 1 diagnosed |
| Belief own life is meaningful, 20460 | Definition: Binary: 0 not diagnosed, 1 diagnosed | / | / | Belief own life is meaningful | Definition: WHOQOL item 6: 1 Not at all, 2 A little, 3 A moderate amount, 4 Very much, 5 An extreme amount |

O, number of outcomes; TRD, treatment resistant depression; MADRS, **Montgomery-Asberg Depression Rating Scale**; DSM, Diagnostic and Statistical Manual of Mental Disorders; MINI, Mini International Neuropsychiatric Interview; BDI, Beck Depression Inventory; WHOQOL, World Health Organization Quality of Life.

**Table S10.** TDA graph characteristics and community detection results for each predictor feature set obtained in UKB. *Robust communities* indicate the communities containing more than 5 samples, considered statistically unreliable and systematically included for subsequent analyses.

|  | **N subjects, P features** | **Graph characteristics** (fixed Filtering with 2-UMAP components and Covering parameters R=15, G=0.80) | **Best Community detection Algorithm** | **Modularity of the partition** | **Total number of communities obtained** | **Numerosity of core samples (100% membership)** |
| --- | --- | --- | --- | --- | --- | --- |
| **G-E-Imaging** | **N=3044**  **P=659+4 covariates** | - **Clustering**: DBSCAN | **Louvain algorithm** | 0.44 | - N=6 total communities extracted - N=4 robust communities | **COM1**:154  **COM2**: 177  **COM3**: 208  **COM4**: 456 |
| **E-Imaging** | **N=3044**  **P=639+4 covariates** | - **Clustering**: DBSCAN | **Louvain algorithm** | 0.43 | - N=5 total communities extracted - N=4 robust communities | **COM1**: 130  **COM2**: 147  **COM3**: 468  **COM4**: 284 |
| **Imaging** | **N=3044**  **P=606+4 covariates** | - **Clustering**: DBSCAN | **Louvain algorithm** | 0.46 | - N=7 total communities extracted - N=4 robust communities | **COM1**: 175  **COM2**: 268  **COM3**: 258  **COM4**: 408 |
| **G-E** | **N=20175**  **P=53 + 4 covariates** | - **Clustering**: HDBSCAN | **Louvain algorithm** | 0.50 | - N=7 total communities extracted - N=4 robust communities | **COM1**: 753  **COM2**: 1089  **COM3**: 2808  **COM4**: 2450 |
| **Environment** | **N=20175**  **P=33 + 4 covariates** | - **Clustering**: HDBSCAN | **Louvain algorithm** | 0.60 | - N=10 total communities extracted - N=4 robust communities | **COM1**: 212  **COM2**: 51  **COM3**: 4084  **COM4**: 8287 |
| **Genes** | **N=20175**  **P=20 + 4 covariates** | - **Clustering**: HDBSCAN | **Louvain algorithm** | 0.50 | - N=5 total communities extracted - N=4 robust communities | **COM1**: 1284  **COM2**: 1059  **COM3**: 1057  **COM4**: 935 |

COM, community; DBSCAN, Density-Based Spatial Clustering of Applications with Noise; HDBSCAN, Hierarchical Density-Based Spatial Clustering of Applications with Noise; N, number of subjects; P, number of features.

**Table S11.** TDA graph characteristics and community detection results for each predictor feature set in GSRD.

|  | **N subjects, P features** | **Graph characteristics**  (fixed Filtering with UMAP 2 components and Covering parameters R=15, G=0.80) | **Best Community detection Algorithm** | **Modularity of the partition** | **Total number of communities obtained** | **Numerosity of core samples (100% membership)** |
| --- | --- | --- | --- | --- | --- | --- |
| **G-E** | **N=1017**  **P=23 + 2 covariates** | - **Clustering**: DBSCAN | **Louvain algorithm** | 0.65 | - N=6 total communities extracted - N=3 robust communities | **COM1**: 223  **COM2**: 255  **COM3**: 229 |
| **Env** | **N=1017**  **P=5 + 2 covariates** | - **Clustering**: DBSCAN | **Louvain algorithm** | 0.97 | - N=45 total communities extracted - N=13 robust communities | **COM1**: 29  **COM2**: 31  **COM3**: 58  **COM4**: 35  **COM5:** 36  **COM6:** 40  **COM7:** 32  **COM8:** 64  **COM9:** 113  **COM10:** 113  **COM11:** 117  **COM12:** 67  **COM12:** 76 |
| **Genes** | **N=1017**  **P=18 + 2 covariates** | - **Clustering**: DBSCAN | **Louvain algorithm** | 0.48 | - N=6 total communities extracted - N=4 robust communities | **COM1**: 100  **COM2**: 49  **COM3**: 117  **COM4**: 123 |

COM, community; DBSCAN, Density-Based Spatial Clustering of Applications with Noise; HDBSCAN, Hierarchical Density-Based Spatial Clustering of Applications with Noise; N, number of subjects; P, number of features.

**Table S12.** TDA graph characteristics and community detection results for each predictor feature set in HSR.

|  | **N subjects, P features** | **Graph characteristics**  (fixed Filtering with UMAP 2 components and Covering parameters R=15, G=0.80) | **Best Community detection Algorithm** | **Modularity of the partition** | **Total number of communities obtained** | **Numerosity of core samples (100% membership)** |
| --- | --- | --- | --- | --- | --- | --- |
| **G-E-Imaging** | **N=71**  **P=375+4 covariates** | - **Clustering**: DBSCAN | **Louvain algorithm** | 0.45 | - N=3 total communities extracted - N=3 robust communities | **COM1**: 26  **COM2**: 9  **COM3**: 13 |
| **E-Imaging** | **N=71**  **P=356+4 covariates** | - **Clustering**: DBSCAN | **Greedy algorithm** | 0.65 | - N=3 total communities extracted - N=3 robust communities | **COM1**: 15  **COM2**: 14  **COM3**: 13 |
| **Imaging** | **N=71**  **P=344+4 covariates** | - **Clustering**: DBSCAN | **Greedy algorithm** | 0.27 | - N=3 total communities extracted - N=2 robust communities | **COM1**: 28  **COM2**: 13 |
| **G-E** | **N=87**  **P=31 + 4 covariates** | - **Clustering**: HDBSCAN | **Louvain algorithm** | 0.44 | - N=5 total communities extracted - N=3 robust communities | **COM1**: 16  **COM2**: 14  **COM3**: 7 |
| **Env** | **N=87**  **P=12 + 4 covariates** | - **Clustering**: HDBSCAN | **Louvain algorithm** | 0.46 | - N=4 total communities extracted - N=4 robust communities | **COM1**: 20  **COM2**: 6  **COM3**: 6  **COM4**: 11 |
| **Genes** | **N=87**  **P=19 + 4 covariates** | - **Clustering**: HDBSCAN | **Louvain algorithm** | 0.54 | - N=8 total communities extracted - N=3 robust communities | **COM1**: 13  **COM2**: 11  **COM3**: 9 |

COM, community; DBSCAN, Density-Based Spatial Clustering of Applications with Noise; HDBSCAN, Hierarchical Density-Based Spatial Clustering of Applications with Noise; N, number of subjects; P, number of features.

**Table S13.** Community health outcome characterization and risk ranking for best predictor feature sets in UKB. *Red colors* highlight **high-risk categories percentage associated with each variable. High risk categories** were defined: (i) for binary outcomes, as the presence of the risk condition; (ii) for ordinal scales, as the set of categories reflecting negative outcomes.

| **Health Outcomes** | **Best predictor** | **Available outcome for each COM** | **Characterization of the outcome** | | | | | **Ranking descending risk** |
| --- | --- | --- | --- | --- | --- | --- | --- | --- |
| TRD | G-E | **C1**:  160  **C2**: 162  **C3**:  439  **C4**: 482 | **C1:** 11.25% **Y**/88.75% N  **C2:** 5.50% **Y**/94.50% N  **C3:** 4.4% **Y**/95.6% N  **C4:** 11.20% **Y**/ 88.8% N | | | | | %yes answer:  C1 (11.25%)  C4 (11.20%)  C2 (5.50%)  C3 (4.4%) |
| Depression for atypical neurovegetative symptoms | G-E | **C1**:  448  **C2**: 634  **C3**:  1829  **C4**: 1602 | **C1:** 8.7% Y/ 91.3% N  **C2:** 3.15% Y/ 96.85%N  **C3:** 5% Y/ 95% N  **C4:** 9% Y/ 91% N | | | | | %yes answer:  C4 (9%)  C1 (8.7%)  C3 (5%)  C2 (3.15%) |
| Depression with anxious features | G-E | **C1**:  448  **C2**: 634  **C3**:  1829  **C4**: 1602 | **C1:** 31.50% Y/68.50% N  **C2:** 22.7% Y/ 77.30% N  **C3:** 26.2% Y/ 73.8% N  **C4:** 33% Y/67% N | | | | | %yes answer:  C4 (33%)  C1 (31.50%)  C3 (26.2%)  C2 (22.7%) |
| N depression episodes | G-E | **C1**:  253  **C2**: 361  **C3**:  923  **C6**: 757 | **C1:** 3, 4; 8.11, 35.2  **C2:** 2, 3; 4.34, 9.64  **C3:** 2, 3; 4.07, 9.78  **C4** 3, 4; 7.92, 23.10  **(Median, iqr; mean, std)** | | | | | Mean and std:  C1 (8.11,35.2)  C4 (7.92, 23.10  C2 (4.34, 9.64)  C3 (4.07, 9.78) |
| Health Satisfaction | G-E | **C1**: 253  **C2**: 361  **C3**: 923  **C4**: 757 |  | **C1** | **C2** | **C3** | **C4** | % total unhappy answer:  C2 (92.5%)  C3 (90.4%)  C1 (81.42%)  C4 (79.39%) |
|  |  |  | **ExtremelyHap** | 3.16% | 0.28% | 0.43% | 2.11% |  |
|  |  |  | **VeryHap** | 3.92% | 1.12% | 1% | 4.2% |  |
|  |  |  | **ModerateHap** | 11.5% | 6.1% | 8.17% | 14.3% |  |
|  |  |  | **ModerateUnHap** | 45.45% | 49% | 51% | 45.7% |  |
|  |  |  | **VeryUnHap** | 31.62% | 38.5% | 34.2% | 29.33% |  |
|  |  |  | **ExtremelyUnHap** | 4.35% | 5% | 5.2% | 4.36% |  |
| Family rel. Satisfaction | G-E | **C1**: 253  **C2**: 361  **C3**: 923  **C4**: 757 |  | **C1** | **C2** | **C3** | **C4** | % total unhappy answer:  C3 (95.4%)  C2 (94.6%)  C1 (92.5%)  C4 (86%) |
|  |  |  | **ExtremelyHap** | 1.6% | 0.3% | 0.2% | 2% |  |
|  |  |  | **VeryHap** | 1.20% | 1.11% | 0.4% | 3% |  |
|  |  |  | **ModerateHap** | 4.7% | 3.90% | 4% | 9% |  |
|  |  |  | **ModerateUnHap** | 29.3% | 28.20% | 24.20% | 36% |  |
|  |  |  | **VeryUnHap** | 41.10% | 45.31% | 51.20% | 37% |  |
|  |  |  | **ExtremelyUnHap** | 22.10% | 21% | 20% | 13% |  |
| Friendship rel. Satisfaction | G-E | **C1**: 253  **C2**: 361  **C3**: 923  **C4**: 757 |  | **C1** | **C2** | **C3** | **C4** | % total unhappy answer:  C2 (96.1%)  C3 (96%)  C1 (95.5%)  C4 (93.1%) |
|  |  |  | **ExtremelyHap** | 0% | 0% | 0% | 0.13% |  |
|  |  |  | **VeryHap** | 0.40% | 0.30% | 0.33% | 1.6% |  |
|  |  |  | **ModerateHap** | 4.15% | 3.6% | 3.4% | 5.2% |  |
|  |  |  | **ModerateUnHap** | 26% | 31.1% | 25% | 35.4% |  |
|  |  |  | **VeryUnHap** | 51% | 51% | 58% | 46.2% |  |
|  |  |  | **ExtremelyUnHap** | 18% | 14% | 13.27% | 11.2% |  |
| Work Job Satisfaction  (0:6 categ) | NS | / | / | | | | | / |
| Duration of worst depression | G-E | **C1**:  647  **C2**: 937  **C3**: 2479  **C4**: 2212 | **C1:** 32% Below3M, 39% Below1Y, 29% Upto1Y  **C2:** 40% Below3M, 41% Below1Y, 19% Upto1Y  **C3:** 42% Below3M, 40% Below1Y, 18% Upto1Y  **C4:** 30% Below3M, 40% Below1Y, 30% Upto1Y | | | | | % Upto1Year answer:  C4 (30%)  C1 (29%)  C2 (19%)  C3 (18%) |
| Frequency of depressed days | G-E | **C1**:  647  **C2**: 937  **C3**: 2479  **C4**: 2212 | **C1:** 4% Less Often, 47% Almosteveryday, 49% Everyday  **C2:** 2% Less Often, 48% Almosteveryday, 50% Everyday  **C3:** 2% Less Often, 46% Almosteveryday, 52% Everyday  **C4:** 2% Less Often, 40% Almosteveryday, 58% Everyday | | | | | % Everyday answer:  C4 (58%)  C3 (52%)  C2 (50%)  C1 (49%) |
| Impact on normal roles | G-E | **C1**:  647  **C2**: 937  **C3**: 2479  **C4**: 2212 | **C1:** 3% Not at all, 15% A little, 82% A lot  **C2:** 1% Not at all, 15.2% A little, 83.8% A lot  **C3:** 2% Not at all, 13% A little, 85% A lot  **C4:** 2.5% Not at all, 13% A little, 84.5% A lot | | | | | % A lot answer:  C3 (85%)  C4 (84.5%)  C2 (83.8%)  C1 (82%) |
| Thought of deadth | G-E | **C1**:  647  **C2**: 937  **C3**: 2479  **C4**: 2212 | **C1:** 62% Y/ 38% N  **C2:** 55% Y/ 45% N  **C3:** 53% Y/ 47% N  **C4:** 64% Y/ 36% N | | | | | % yes answer:  C4 (64%)  C1 (62%)  C2 (55%)  C3 (53%) |
| Ever self harmed | G-E | **C1**:  647  **C2**: 937  **C3**: 2479  **C4**: 2212 | **C1:** 12% Y/ 88% N  **C2:** 7% Y/ 93% N  **C3:** 3% Y/ 97% N  **C4:** 13% Y/ 87% N | | | | | % yes answer:  C4 (13% )  C1 (12%)  C2 (7%)  C3 (3%) |
| Belief own life is meaningful | G-E | **C1**:  647  **C2**: 937  **C3**: 2479  **C4**: 2212 |  | **C2** | **C2** | **C3** | **C4** | % moderate, very much, extreme answers:  C3 (94.03%)  C2 (91.6%)  C1 (86.5%)  C4 (83.2%) |
|  |  |  | **Not at all** | 3.4% | 1.28% | 0.97% | 5% |  |
|  |  |  | **A little** | 10.2% | 7.15% | 5% | 13% |  |
|  |  |  | **Moderateamount** | 30.6% | 30.6% | 26.5% | 35% |  |
|  |  |  | **Very much** | 44% | 50.5% | 54.3% | 40.8% |  |
|  |  |  | **Extremeamount** | 11.8% | 10.47% | 13.23% | 8.2% |  |
| Depression related to stress or traumatic events | G-E-Imaging | **C1**: 134  **C2**: 158  **C3**: 191  **C4**: 414 | **C1:**   81% Y/ 19% N  **C2:** 65% Y/ 35% N  **C3:**  66% Y/ 34% N  **C4:** 76% Y/ 24% N | | | | | % yes answer:  C1 (81% )  C4 (76% )  C3 (66% )  C2 (65% ) |
| Cancer diagnosed | E-Imaging | **C1**: 130  **C2**: 147  **C3**:  468  **C4**: 184 | **C1:** 8.5% Y/ 91.5% N  **C2:** 9% Y/ 91% N  **C3:** 3.2% Y/ 96.8% N  **C4:** 3.5% Y/ 96.5% N | | | | | % yes answer:  C2 (9% )  C1 (8.5% )  C4 (3.5% )  C3 (3.2% ) |
| Diabetes diagnosed | Genes | **C1**: 1281  **C2**: 1056  **C3**:  1054  **C4:** 931 | **C1:** 1.4% Y/ 98.6 % N  **C2:** 4% Y/ 96% N  **C3:** 4.4% Y/ 95.6% N  **C4:** 3.3% Y/ 96.7% N | | | | | % yes answer:  C3 (4.4%)  C2 (4% )  C4 (3.3%)  C1 (1.4%) |
| Vascular problems diagnosed | Imaging | **C1:** 175  **C2:** 268  **C3:** 258  **C4:** 408 | **C1:** 82% N, 2% HBP, 16% HBM,stroke,angina  **C2:** 66% N, 1% HBP, 33% HBM, stroke,angina  **C3:** 88% N, 2% HBP, 10% HBM, stroke,angina  **C4:** 82% N, 0.5% HBP, 17,5% HBM, stroke,angina | | | | | % yes answer:  C2 (33%)  C4 (17.5%)  C1 (16%)  C3 (10%) |

G-E-Imaging, genetic-environment; G-E, genetic-environment; HBM, high-blood pressure; C, Community; TRD, treatment-resistant depression.

**Table S14.** Community characterization of top-ranked features profiles and socio-demographic factors across best predictor feature sets in UKB.

| **Best predictor** | **Socio-demographic characteristics** | **First 10 Top-ranked features** | **COM1** | **COM2** | **COM3** | **COM4** | **Ranking descending order** |
| --- | --- | --- | --- | --- | --- | --- | --- |
| **G-E** |  |  | **COM1: 753 sample** | **COM2: 2450 sample** | **COM3: 2808 sample** | **COM4: 1089 sample** |  |
|  |  | Alcohol intake frequency (0-3 levels) | 0:9,4%  1:25.4%  2: 44.2%  3: 21% | 0:10.7%  1: 32.4%  2: 55.4%  3: 1.5% | 0: 5.2%  1: 25%  2: 69.8%  3: 0% | 0: 0.6%  1: 2.7%  2: 7.2%  3: 89.4% | Ranking descending based on %almost daily answer:  **COM4** (89.44%)  COM1  COM2  COM3 |
|  |  | Adulthood stress (continuous) | Mean: 6.55  std: 2.92  Median: 6  IQR: 4 | Mean: 3.88  std: 2  Median: 4  IQR: 2 | Mean: 0.73  std: 1.2  Median: 0  IQR: 1 | Mean: 1.40  std: 1.80  Median: 1  IQR: 2 | Ranking descending based on mean and std:  **COM1 (6.55)**  COM2  COM4  COM3 |
|  |  | Belittlement by partner or ex-partner as an adult (0-5 levels) | 0: 71.3%  1: 7%  2: 9.7%  3: 3%  4: 8.9% | 0: 28.6%  1: 13.9%  2: 40.2%  3: 10.1%  4: 7.1% | 0: 94%  1: 6%  2: 0.1%  3: 0%  4: 0% | 0: 79.9%  1: 9.6%  2: 8.8%  3: 1.4%  4: 0.3% | Ranking descending based on %often answers (corresponding to 3+4 answer):  **COM2** (17.22%)  COM1 (11.95%)  COM4 (1.65%)  COM3 0% |
|  |  | Someone to take to doctor when needed as a child (0-5 levels) | 0: 14.7%  1: 3.9%  2: 6.6%  3: 12.3%  4: 62.4% | 0: 0.5%  1: 1.4%  2: 7%  3: 14.5%  4: 76.7% | 0: 0%  1: 0%  2: 0.1%  3: 4.8%  4: 95.1% | 0: 0%  1: 0.2%  2: 1.5%  3: 7.9%  4: 90.5% | Ranking descending based on %often answers (corresponding to 3+4 answer:  **COM3** (99.89%)  COM4 (98.35%)  COM2 (91.18%)  COM1 (74.77%) |
|  |  | Felt hated by family member as a child (0-5 levels) | 0: 77.7%  1: 5.2%  2: 8.2%  3: 3%  4: 5.8% | 0:63.2%  1: 10.1%  2: 16.7%  3: 5.9%  4: 4.1**%** | 0: 97%  1: 2.9%  2: 0.1%  3: 0%  4: 0% | 0: 89.3%  1: 5.6%  2: 4.4%  3: 0.4%  4: 0.4% | Ranking descending based on %often answers (corresponding to 3+4 answer :  **COM2** (10.04%)  COM1 (8.90%)  COM4 (0.73%)  COM3 (0%) |
|  |  | Felt loved as a child (0-5 levels) | 0: 5.3%  1: 10.9%  2: 19.6%  3: 21.2%  4: 42.9% | 0: 3.8%  1: 12.5%  2: 27.9%  3: 21.8%  4: 34.0% | 0: 0.1%  1: 0.8%  2: 7.6%  3: 22.5%  4: 69% | 0: 0.9%  1: 3.2%  2: 16.1%  3: 23.8%  4: 56.0% | Ranking ascending based on %often answers (corresponding to 3+4 answer :  **COM2** (66.80%)  COM1 (64.14%)  COM4 (79.80%)  COM3 (91.56%) |
|  |  | Physical violence by partner or ex-partner as an adult (0-5 levels) | 0: 81.8%  1: 5.4%  2: 5.6%  3: 1.6%  4: 5.6% | 0: 74.2%  1: 15%  2: 10%  3: 0%  4: 0% | 0: 97.9%  1: 2%  2: 0%  3: 0%  4:0 % | 0: 89.2%  1: 6.4%  2: 4.2%  3: 0.1%  4: 0.1% | Ranking descending based on %often answers (corresponding to 3+4 answer:  **COM1** (7.17%)  COM2 (0.78%)  COM4 (0.18%)  COM3 0% |
|  |  | Physically abused by family as a child (0-5 levels) | 0: 76.5%  1: 9%  2: 10.8%  3: 1.5%  4: 2.3% | 0: 68.4%  1: 14%  2: 14.2%  3: 2.3%  4: 1.1% | 0: 94.3%  1: 5.7%  2: 0.1%  3: 0%  4: 0% | 0: 84.4%  1: 10.4%  2: 4.9%  3: 0.4%  4: 0% | Ranking descending based on %often answers (corresponding to 3+4 answer :  **COM1** (3.72%)  COM2 (3.39%)  COM4 (0.37%)  COM3 (0%) |
|  | Age |  | Mean: 55.84  std: 7.60  Median: 56  IQR: 12 | Mean: 53.31  std: 7.56  Median: 53  IQR: 12 | Mean: 55  std: 7.28  Median: 55  IQR: 12 | Mean: 56.46  std: 7.39  Median: 57  IQR: 11 |  |
|  | Sex |  | 0: 65.5%  1: 37.5% | 0: 74.6%  1: 25.4% | 0: 67.2%  1: 32.8% | 0: 53.7%  1: 46.3% |  |
|  | Ethnicity |  | 0: 1.3%  1: 98.7% | 0: 0.9%  1: 99.1% | 0: 0.2%  1: 99.8% | 0: 0.2%  1: 99.8% |  |
|  | Antidepressant |  | 0: 84.9%  1: 15.1% | 0: 80.9%  1: 19.1% | 0: 89.4%  1: 10.6% | 0: 88.1%  1: 11.9% |  |
| **G-E-Imaging** |  |  | **COM1: 154 sample** | **COM2: 177 sample** | **COM3: 208 sample** | **COM4: 456 sample** |  |
|  |  | Area of Superiorfrontal - Right | Mean: 8010.416  std: 654.124  Median: 7957  IQR: 835 | Mean:9719  std:745  Median: 9672  IQR: 866 | Mean: 9691  std: 857  Median: 9705  IQR: 1105 | Mean: 7908  std: 621  Median: 7912  IQR: 825.5 | Descending order based on mean and std:  **COM2**  COM3  COM1  COM4 |
|  |  | Area of Superiorfrontal - Left | Mean: 7096.104  std: 585.828  Median: 7101.500  IQR: 777.500 | Mean: 8458.390  std: 652.435  Median: 8426.000  IQR: 746 | Mean: 8493.524  std: 717.308  Median: 8454.500  IQR: 1027.500 | Mean: 6957.950  std: 561.023  Median: 6975.500  IQR: 751.250 | Descending order:  **COM3**  COM2  COM1  COM4 |
|  |  | Area of Superiortemporal - Right | Mean: 4395.435  std: 299.978  Median: 4428.000  IQR: 351.750 | Mean: 5112.322  std: 336.195  Median: 5111  IQR: 427 | Mean: 5087.928  std: 356.269  Median: 5066  IQR: 472.250 | Mean: 4323.401  std: 315.153  Median: 4324.500  IQR: 426.250 | Descending order based on mean and std:  **COM2**  COM3  COM1  COM4 |
|  |  | Mean RD in SuperiorCoronaRadiata on FA skeleton - Left | Mean:5.55e-04  std: 2.23e-05  Median:0.00055  IQR: 2.84e-05 | Mean:5.37e-04  std: 2.61e-05  Median: 0.00054  IQR:3.3e-05 | Mean:4.86e.04  std: 1.87e-05  Median: 0.00049  IQR:2.55e-05 | Mean:4.96e-04  std:1.82e-05  Median: 0.00049  IQR:2.40e-05 | Descending order based on mean and std:  **COM1**  COM2  COM4  COM3 |
|  |  | Area of Superiortemporal - Left | Mean: 4682.903  std: 338.435  Median: 4629.500  IQR: 414.500 | Mean: 5459.780  std: 424.663  Median: 5445  IQR: 523 | Mean: 5469.519  std: 436.329  Median: 5437.500  IQR: 578 | Mean: 4585.327  std: 343.706  Median: 4569  IQR: 455 | Descending order based on mean and std:  **COM3**  COM2  COM1  COM4 |
|  |  | Area of Lateralorbitofrontal - Left | Mean: 2720.955  std: 218.692  Median: 2731  IQR: 327 | Mean: 3215.989  std: 229.421  Median: 3230  IQR: 328 | Mean: 3206.274  std: 243.091  Median: 3197  IQR: 320.500 | Mean: 2711.884  std: 213.403  Median: 2729.500  IQR: 266 | Descending order based on mean and std:  **COM2**  COM3  COM1  COM4 |
|  |  | Mean RD in SuperiorCoronaRadiata on FA skeleton - Right | Mean: 5.54e-04  std: 2.42e-05  Median: 5.50e-04  IQR: 2.96e-05 | Mean: 5.35e-04  std: 2.34e-05  Median: 5.35e-04  IQR: 3.30e-05 | Mean: 4.87e-04  std: 1.96e-05  Median: 4.87e-04  IQR: 2.69e-05 | Mean: 4.98e-04  std: 1.83e-05  Median: 4.99e-04  IQR: 2.65e-05 | Descending order based on mean and std:  **COM1**  COM2  COM4  COM3 |
|  |  | Mean RD in PosteriorCoronaRadiata on FA skeleton - Left | Mean: 6.29e-04  std: 4.76e-05  Median: 6.20e-04  IQR: 4.54e-05 | Mean: 6.03e-04  std: 3.77e-05  Median: 6.01e-04  IQR: 3.55e-05 | Mean: 5.40e-04  std: 2.49e-05  Median: 5.42e-04  IQR: 3.26e-05 | Mean: 5.45e-04  std: 2.46e-05  Median: 5.44e-04  IQR: 3.05e-05 | Descending order based on mean and std:  **COM1**  COM2  COM4  COM3 |
|  |  | Area of Lateralorbitofrontal - Right | Mean: 2711.299  std: 227.675  Median: 2686.500  IQR: 306.500 | Mean: 3200.582  std: 250.613  Median: 3191  IQR: 327 | Mean: 3230.418  std: 253.706  Median: 3228.500  IQR: 388 | Mean: 2698.042  std: 223.039  Median: 2700.500  IQR: 317.750 | Descending order based on mean and std:  **COM3**  COM2  COM1  COM4 |
|  |  | Mean RD in PosteriorCoronaRadiata on FA skeleton - Right | Mean: 6.35e-04  std: 4.15e-05  Median: 6.27e-04  IQR: 5.10e-05 | Mean: 6.09e-04  std: 4.24e-05  Median: 6.08e-04  IQR: 5.25e-05 | Mean: 5.46e-04  std: 2.51e-05  Median: 5.47e-04  IQR: 3.13e-05 | Mean: 5.48e-04  std: 2.44e-05  Median: 5.48e-04  IQR: 2.95e-05 | Descending order based on mean and std:  **COM1**  COM2  COM4  COM3 |
|  | Age |  | Mean: 57.95  std: 5.97  Median: 59  IQR: 8 | Mean: 56.85  std: 6.02  Median: 57  IQR: 9 | Mean: 50.35  std: 6.16  Median: 50  IQR: 10 | Mean: 52.14  std: 6.76  Median: 52  IQR: 10 |  |
|  | Sex |  | 0: 89%  1: 11% | 0: 78.5%  1: 21.5% | 0: 71.6%  1: 18.4% | 0: 93.6%  1: 6.4% |  |
|  | Ethnicity |  | 0: 0  1: 100% | 0: 0  1: 100% | 0: 1%  1: 99% | 0: 1.3%  1: 98.7% |  |
|  | Antidepressant |  | 0: 87%  1: 13% | 0: 87%  1: 13% | 0: 88.9%  1: 11.1% | 0: 86.2%  1: 13.8% |  |
| **E-Imaging** |  |  | **COM1: 130 samples** | **COM2: 147 samples** | **COM3: 468 samples** | **COM4: 284 samples** |  |
|  |  | Area of Superiorfrontal - Left | Mean: 6944.39  std: 574.50  Median: 6917.50  IQR: 810.75 | Mean: 8207.16  std: 651.19  Median: 8199  IQR: 829 | Mean: 7074.86  std: 539.98  Median: 7069.50  IQR: 671 | Mean: 8593.51  std: 687.10  Median: 8508.50  IQR: 948 | Descending order based on mean and std:  COM4  COM3  COM2  **COM1** |
|  |  | Area of Rostralanteriorcingulate - Left | Mean: 997.34  std: 138.35  Median: 1004.50  IQR: 169.75 | Mean: 1280.37  std: 158.85  Median: 1285.00  IQR: 221 | Mean: 1038.51  std: 137.04  Median: 1035  IQR: 178.25 | Mean: 1371.76  std: 164.16  Median: 1357  IQR: 219.75 | Descending order based on mean and std:  COM4  COM3  COM2  **COM1** |
|  |  | Area of Middletemporal - Right | Mean: 3708.50  std: 343.83  Median: 3715.00  IQR: 448.75 | Mean: 4335.37  std: 351.88  Median: 4333  IQR: 433.50 | Mean: 3810  std: 306.82  Median: 3809.50  IQR: 392 | Mean: 4593.03  std: 430.95  Median: 4540.00  IQR: 552.25 | Descending order based on mean and std:  COM4  COM2  COM3  **COM1** |
|  |  | Area of Precentral - Left | Mean: 4274.45  std: 343.86  Median: 4235.50  IQR: 374.75 | Mean: 4952.63  Std: 406.65  Median: 4930.00  IQR: 526.00 | Mean: 4331.64  Std: 317.56  Median: 4321.50  IQR: 422.50 | Mean: 5122.06  Std: 412.10  Median: 5100.50  IQR: 525.50 | Descending order based on mean and std:  COM4  COM2  COM3  **COM1** |
|  |  | Area of Precentral – Right | Mean: 4165.15  Std: 381.97  Median: 4160.50  IQR: 472.75 | Mean: 4840.98  Std: 396.90  Median: 4797.00  IQR: 565.50 | Mean: 4209.71  Std: 323.18  Median: 4191.00  IQR: 411.25 | Mean: 4977.39  Std: 419.94  Median: 4966.00  IQR: 551.00 | Descending order based on mean and std:  COM4  COM2  COM3  **COM1** |
|  |  | Area of Postcentral – Left | Mean: 4187.25  Std: 341.52  Median: 4177.00  IQR: 438.50 | Mean: 4843.67  Std: 399.75  Median: 4802.00  IQR: 537.50 | Mean: 4277.02  Std: 343.33  Median: 4270.00  IQR: 449.50 | Mean: 5087.01  Std: 453.72  Median: 5041.00  IQR: 574.50 | Descending order based on mean and std:  COM4  COM2  COM3  **COM1** |
|  |  | Area of Postcentral – Right | Mean: 3959.22  Std: 334.47  Median: 3940.00  IQR: 468.75 | Mean: 4636.11  Std: 415.26  Median: 4594.00  IQR: 557.00 | Mean: 4057.28  Std: 339.06  Median: 4047.50  IQR: 429.25 | Mean: 4838.39  Std: 451.13  Median: 4793.00  IQR: 572.50 | Descending order based on mean and std:  COM4  COM2  COM3  **COM1** |
|  |  | Area of Medialorbitofrontal – Right | Mean: 1375.85  Std: 128.82  Median: 1384.00  IQR: 174.25 | Mean: 1586.35  Std: 127.49  Median: 1594.00  IQR: 166.50 | Mean: 1405.83  Std: 115.96  Median: 1406.00  IQR: 148.50 | Mean: 1662.54  Std: 134.65  Median: 1656.00  IQR: 169.50 | Descending order based on mean and std:  COM4  COM2  COM3  **COM1** |
|  |  | Area of Rostralmiddlefrontal – Left | Mean**:** 3459.54  Std: 363.70  Median: 3454.50  IQR: 454.50 | Mean: 4192.27  Std: 484.70  Median: 4163.00  IQR: 591.00 | Mean: 3581.31  Std: 373.64  Median: 3557.50  IQR: 476.25 | Mean: 4466.73  Std: 513.70  Median: 4404.50  IQR: 707.50 | Descending order based on mean and std:  COM4  COM2  COM3  **COM1** |
|  |  | Area of Rostralmiddlefrontal - Right | Mean: **3585.85**  Std: 422.20  Median: 3518.50  IQR: 487.25 | Mean: 4310.76  Std: 516.23  Median: 4270.00  IQR: 623.50 | Mean: 3702.08  Std: 413.70  Median: 3669.00  IQR: 532.75 | Mean: 4634.65  Std: 485.09  Median: 4627.00  IQR: 599.75 | Descending order based on mean and std:  COM4  COM2  COM3  **COM1** |
|  | Age |  | Mean: 56.73  std: 6.43  Median: 57  IQR: 8.75 | Mean: 58.50  std: 5.89  Median: 59  IQR: 8 | Mean: 50.36  std: 6.21  Median: 50  IQR: 10 | Mean: 50.17  std: 6.09  Median: 50  IQR: 10 |  |
|  | Sex |  | 0: 90.8%  1: 9.2% | 0: 72.1%  1: 27.9% | 0: 90.2%  1: 9.8% | 0: 77.5%  1: 22.5% |  |
|  | Ethnicity |  | 0: 0%  1: 100% | 0: 0%  1: 100% | 0: 0.6%  1: 99.4% | 0: 0.4%  1: 99.6% |  |
|  | Antidepressant Medication |  | 0: 88.5%  1: 11.5% | 0: 89.9%  1: 10.2% | 0: 88%  1: 12% | 0:89.8%  1: 10.2% |  |
| **Genes** |  |  | **COM1: 1284 sample** | **COM2: 1059 sample** | **COM3: 1057 sample** | **COM4: 935** |  |
|  |  | PRS ADHD | Mean: -0.436  Std: 0.745  Median: -0.449  IQR: 1.020 | Mean: **-0.535**  Std: 0.794  Median: -0.520  IQR: 1.004 | Mean: 0.356  Std: 0.796  Median: 0.373  IQR: 1.064 | Mean: 0.648  Std: 0.773  Median: 0.653  IQR: 1.002 | Descending order based on mean and std:  **COM2**  COM1  COM3  COM4 |
|  |  | PRS AUTISM | Mean: 0.0473  Std: 0.756  Median: 0.0606  IQR: 0.995 | Mean: **-0.554**  Std: 0.781  Median: -0.583  IQR: 1.061 | Mean: 0.114  Std: 0.861  Median: 0.101  IQR: 1.171 | Mean: 0.800  Std: 0.798  Median: 0.852  IQR: 1.084 | Descending order based on mean and std:  **COM2**  COM1  COM3  COM4 |
|  |  | PRS Anorexia Nervosa | Mean: 0.214  Std: 0.808  Median: 0.197  IQR: 1.150 | Mean: -0.508  Std: 0.835  Median: -0.525  IQR: 1.160 | Mean: -0.113  Std: 0.852  Median: -0.105  IQR: 1.218 | Mean: 0.530  Std: 0.802  Median: 0.500  IQR: 1.089 | Descending order based on mean and std:  **COM2**  COM3  COM1  COM4 |
|  | Age |  | Mean: 55.89  std: 7.16  Median: 57  IQR: 10 | Mean: 54.03  std: 7.25  Median: 55  IQR: 12 | Mean: 54.06  std: 7.14  Median: 54  IQR: 12 | Mean: 55.03  std: 7.06  Median: 55  IQR: 11 |  |
|  | Sex |  | 0: 69.3%  1: 30.7% | 0: 69.2%  1: 30.8% | 0: 68.3%  1: 31.7% | 0: 68.8%  1: 31.1% |  |
|  | Ethnicity |  | 0: 0.2%  1: 99.8% | 0: 0.1%  1: 99.9% | 0: 0.4%  1: 99,6% | 0: 0.5%  1: 99.5% |  |
|  | Antidepressant |  | 0: 92.4%  1: 7.6% | 0: 89.3%  1: 10.7% | 0: 85.6%  1: 14.4% | 0: 90.2%  1: 9.8% |  |
| **Imaging** |  |  | **COM1: 175 sample** | **COM2: 258 sample** | **COM3: 268 sample** | **COM4: 408 sample** |  |
|  |  | Area of Superiorfrontal – Left | Mean: 6839.54  Std: 539.83  Median: 6871.00  IQR: 757.00 | Mean: 7093.49  Std: 474.01  Median: 7094.00  IQR: 602.00 | Mean: 7851.22  Std: 723.50  Median: 7825.00  IQR: 899.25 | Mean: 8565.67  Std: 701.75  Median: 8495.00  IQR: 933.75 | Descending order based on mean and std:  COM4  COM3  **COM2**  **COM1** |
|  |  | Area of Superiorfrontal - Right | Mean: 7773.48  Std: 613.58  Median: 7779.00  IQR: 778.00 | Mean: 8109.89  Std: 628.08  Median: 8061.50  IQR: 828.00 | Mean: 8933.09  Std: 835.79  Median: 8912.00  IQR: 1085.50 | Mean: 9818.17  Std: 818.99  Median: 9737.00  IQR: 1125.25 | Descending order based on mean and std:  COM4  COM3  **COM2**  **COM1** |
|  |  | Area of Superiortemporal - Left | Mean: 4527.40  Std: 327.89  Median: 4479.00  IQR: 398.50 | Mean: 4658.59  Std: 319.82  Median: 4656.00  IQR: 469.75 | Mean: 5090.16  Std: 448.81  Median: 5060.50  IQR: 567.00 | Mean: 5587.41  Std: 457.14  Median: 5552.50  IQR: 644.25 | Descending order based on mean and std:  COM4  COM3  **COM2**  **COM1** |
|  |  | Mean RD in SuperiorCoronaRadiata on FA skeleton - Left | Mean: 5.28e-04  Std: 1.83e-05  Median: 5.26e-04  IQR: 2.32e-05 | Mean: 4.82e-04  Std: 1.60e-05  Median: 4.83e-04  IQR: 2.19e-05 | Mean: 5.42e-04  Std: 2.99e-05  Median: 5.40e-04  IQR: 3.81e-05 | Mean: 5.02e-04  Std: 2.02e-05  Median: 5.03e-04  IQR: 2.50e-05 | Descending order based on mean and std:  **COM2**  COM4  COM1  COM3 |
|  |  | Area of Superiortemporal - Right | Mean: 4268.40  Std: 299.45  Median: 4259.00  IQR: 416.50 | Mean: 4399.78  Std: 294.00  Median: 4404.50  IQR: 421.75 | Mean: 4786.92  Std: 388.13  Median: 4771.50  IQR: 507.25 | Mean: 5173.50  Std: 396.19  Median: 5158.00  IQR: 544.75 | Descending order based on mean and std:  COM4  COM3  **COM2**  **COM1** |
|  |  | Mean RD in SuperiorFronto_occipitalFasciculus on FA skeleton - Left | Mean: 5.39e-04  Std: 3.83e-05  Median: 5.32e-04  IQR: 4.53e-05 | Media: 4.85e-04  Std: 3.36e-05  Median: 4.85e-04  IQR: 3.89e-05 | Mean: 6.49e-04  Std: 9.62e-05  Median: 6.34e-04  IQR: 1.17e-04 | Mean: 5.06e-04  Std: 4.30e-05  Median: 5.01e-04  IQR: 4.18e-05 | Descending order based on mean and std:  **COM2**  COM4  COM1  COM3 |
|  |  | Area of Lateral Orbitofrontal - Left | Mean: 2655.74  Std: 214.59  Median: 2644.00  IQR: 274.50 | Mean: 2777.86  Std: 217.38  Median: 2782.00  IQR: 261.75: | Mean: 2977.66  Std: 270.53  Median: 2993.00  IQR: 373.50 | Mean: 3257.92  Std: 241.67  Median: 3240.50  IQR: 325.25 | Descending order based on mean and std:  COM4  COM3  **COM2**  **COM1** |
|  |  | Mean RD in SuperiorCoronaRadiata on FA skeleton - Right | Mean: 5.28e-04  Std: 1.99e-05  Median: 5.26e-04  IQR: 2.68e-05 | Mean: 4.84e-04  Std: 1.76e-05  Median: 4.81e-04  IQR: 2.10e-05 | Mean: 5.42e-04  Std: 2.92e-05  Median: 5.40e-04  IQR: 3.73e-05 | Mean: 5.03e-04  Std: 2.02e-05  Median: 5.04e-04  IQR: 2.86e-05 | Descending order based on mean and std:  **COM2**  COM4  COM1  COM3 |
|  |  | Area of Lateral Orbitofrontal - Right | Mean: 2658.60  Std: 237.31  Median: 2636.00  IQR: 321.00 | Mean: 2788.61  Std: 208.16  Median: 2784.00  IQR: 293.00 | Mean: 2992.50  Std: 278.24  Median: 2966.50  IQR: 384.25 | Mean: 3293.18  Std: 261.72  Median: 3284.50  IQR: 338.75 | Descending order based on mean and std:  COM4  COM3  **COM2**  **COM1** |
|  |  | Area of Rostralanteriorcingulate - Left | Mean: 992.46  Std: 125.48  Median: 990.00  IQR: 160.50 | Mean: 1044.11  Std: 138.97  Median: 1037.50  IQR: 177.00 | Mean: 1199.51  Std: 173.27  Median: 1200.00  IQR: 224.00 | Mean: 1361.62  Std: 163.47  Median: 1348.50  IQR: 216.00 | Descending order based on mean and std:  COM4  COM3  **COM2**  **COM1** |
|  | Age |  | Mean: 55.09  std: 6.70  Median: 55  IQR: 10 | Mean: 49.07  std: 5.98  Median: 48  IQR: 9 | Mean: 59.91  std: 5.54  Median: 61  IQR: 7 | Mean: 50.58  std: 6.30  Median: 50  IQR: 10 |  |
|  | Sex |  | 0: 96.6%  1: 3.4% | 0: 90.3%  1: 9.7% | 0: 50.7%  1: 49.3% | 0: 77%  1: 23% |  |
|  | Ethnicity |  | 0: 1.7%  1: 98.3% | 0: 0.4%  1: 99.6% | 0: 0%  1: 100% | 0:99.5%  1: 0.5% |  |
|  | Antidepressant |  | 0: 87.4%  1: 12.6% | 0: 87.2%  1: 12.8% | 0: 86.9%  1: 13.1% | 0: 89.5%  1: 10.5% |  |

COM, community; Std, standard deviation; IQR, interquartile range; PRS, Polygenetic Risk scores; TRD, treatment resistant depression; RD, radial diffusivity; FA, fractional anisotropy; ADHD, attention deficit hyperactivity disorder; Std, standard deviation; IQR, interquartile range;

**References Supplementary**

Fabbri, C. et al. Genetic and clinical characteristics of treatment-resistant depression using primary care records in two UK cohorts. Mol Psychiatry 26, 3363–3373 (2021).

Fry, A. et al. Comparison of Sociodemographic and Health-Related Characteristics of UKB Participants With Those of the General Population. American Journal of Epidemiology 186, 1026–1034 (2017).

Davis, K. A. S. et al. Mental health in UKB – development, implementation and results from an online questionnaire completed by 157 366 participants: a reanalysis. BJPsych open 6, e18 (2020).

Alfaro-Almagro, F. *et al.* Image processing and Quality Control for the first 10,000 brain imaging datasets from UKB. *NeuroImage* **166**, 400–424 (2018).

Jiang, R. *et al.* A functional connectome signature of blood pressure in &gt;30 000 participants from the UKB. *Cardiovasc. Res.* **119**, 1427–1440 (2023).

Hariri, A. R., Tessitore, A., Mattay, V. S., Fera, F. & Weinberger, D. R. The Amygdala Response to Emotional Stimuli: A Comparison of Faces and Scenes. *NeuroImage* **17**, 317–323 (2002).

Bycroft, C. *et al.* The UKB resource with deep phenotyping and genomic data. *Nature* **562**, 203–209 (2018).

Manichaikul, A. *et al.* Robust relationship inference in genome-wide association studies. *Bioinformatics* **26**, 2867–2873 (2010).

The International Consortium of Blood Pressure (ICBP) 1000G Analyses *et al.* Genome-wide association analysis identifies novel blood pressure loci and offers biological insights into cardiovascular risk. *Nat. Genet.* **49**, 403–415 (2017).

the Haplotype Reference Consortium. A reference panel of 64,976 haplotypes for genotype imputation. *Nat. Genet.* **48**, 1279–1283 (2016).

The UK10K Consortium *et al.* The UK10K project identifies rare variants in health and disease. *Nature* **526**, 82–90 (2015).

Kautzky A, Bartova L, Dold M, Souery D, Montgomery S, Zohar J, Mendlewicz J, Fabbri C, Serretti A, Tretiakov E, Rujescu D, Harkany T, Kasper S. Symptom networks in major depressive disorder and treatment response: special focus on TRD. Eur Psychiatry. 2025 May 27;68(1):e61. doi: 10.1192/j.eurpsy.2025.2454. PMID: 40420429; PMCID: PMC12260719.

Purcell S, Neale B, Todd-Brown K, Thomas L, Ferreira MA, Bender D, Maller J, Sklar P, de Bakker PI, Daly MJ, Sham PC. PLINK: a tool set for whole-genome association and population-based linkage analyses. Am J Hum Genet. 2007 Sep;81(3):559-75. doi: 10.1086/519795. Epub 2007 Jul 25. PMID: 17701901; PMCID: PMC1950838.

Anderson, C. A., Pettersson, F. H., Clarke, G. M., Cardon, L. R., Morris, A. P., & Zondervan, K. T. (2010). Data quality control in genetic case-control association studies. Nature protocols, 5(9), 1564–1573.

Bernstein, D. P., Stein, J. A., Newcomb, M. D., Walker, E., Pogge, D., Ahluvalia, T., ... & Zule, W. (2003). Development and validation of a brief screening version of the Childhood Trauma Questionnaire. *Child abuse & neglect*, *27*(2), 169-190.

Whoqol Group. (1998). Development of the World Health Organization WHOQOL-BREF quality of life assessment. *Psychological medicine*, *28*(3), 551-558.

Reynolds, W. M., & Gould, J. W. (1981). A psychometric investigation of the standard and short form Beck Depression Inventory. *Journal of consulting and clinical psychology*, *49*(2), 306.

Holmes, T. H., & Rahe, R. H. (1967). Schedule of recent experience. *Marriage*, *10*, 50.

Thase ME, Greenhouse JB, Frank E, Reynolds CF 3rd, Pilkonis PA, Hurley K, Grochocinski V, Kupfer DJ. Treatment of major depression with psychotherapy or psychotherapy-pharmacotherapy combinations. Arch Gen Psychiatry. 1997 Nov;54(11):1009-15.
